# Longitudinal antibody profiling after dengue reveals distinct dynamics by antibody specificity over 18 months

**DOI:** 10.1101/2025.08.11.25333449

**Authors:** Sandra Bos, Tulika Singh, José Victor Zambrana, Elias Duarte, Reinaldo Mercado-Hernandez, Julia Huffaker, Aaron Graber, Angel Balmaseda, Eva Harris

## Abstract

The four dengue virus serotypes (DENV1-4) co-circulate worldwide, posing major challenges for vaccine development. One key issue is that certain levels and subsets of cross-reactive antibodies can enhance disease during subsequent infection with a different DENV serotype. We defined the magnitude and kinetics of 84 antiviral antibody subsets (by isotype, subclass, antigen, and cross-reactivity) after primary versus secondary dengue, using longitudinal samples collected <1, 3, 6 and 18 months post-symptom onset from a pediatric hospital study in Nicaragua. Interestingly, we found that post-primary infection, cross-reactive IgG antibodies against the envelope protein rise, not wane, over time. Antibody kinetics varied by specificity as measured by infecting serotype versus cross-reactive subsets, viral antigen, and subdomain of a single antigen. Further, substantial seropositivity of IgA, IgM, and IgG3 at 18 months post-infection was observed. These findings highlight several novel conceptual insights into flavivirus immunity and disease risk and have implications for vaccine design and serodiagnosis.

## Introduction

Dengue virus serotypes 1-4 (DENV1-4) cause febrile illness with intense body pain, occasionally progressing to severe complications including shock.^1^ More than half the world’s population is at risk, with well over 100 million infections annually.^2^ DENV transmission zones are expanding with climate change beyond the tropics and sub-tropics.^3^ Addressing dengue is urgent due to the lack of therapies, inconsistent mosquito control, and limited vaccine availability.^4^ Understanding immune responses is critical for vaccine development.

Antibodies can either protect against or enhance DENV infections.^5–7^ Primary DENV infection is thought to induce lifelong immunity against the infecting serotype but generates cross-reactive (XR) IgG antibodies, which can exacerbate infection with distinct serotypes via antibody-dependent enhancement (ADE).^8,9^ ADE occurs when these antibodies facilitate viral entry into immune cells, leading to increased viral replication, immune activation, and disease severity.^10–12^ Related flaviviruses, such as Zika virus (ZIKV), also produce XR antibodies that can increase the risk of severe dengue during subsequent infections.^13–17^ In contrast, post-secondary DENV infections generally confer broader, protective immunity through higher-quality XR antibodies, although the durability of these responses remains unclear.^18–21^

Current evidence supports only short-term (3–18 months) cross-protection after primary infection.^22–26^ The classical hypothesis underlying severe secondary dengue posits that XR DENV antibodies wane to enhancing levels following primary infection but remain elevated and protective after secondary infection. However, recent studies have found stable long-term antibody levels post-primary infection and slow antibody waning post-secondary infection.^16,27–29^ Thus, antibody waning versus stability post-primary infection remains controversial.

The antigenic relatedness between flaviviruses varies significantly across viral proteins due to differences in their conservation and structure.^30–34^ The envelope (E) protein, forming dimers on the virus surface, elicits most antibodies, with XR dimer-specific neutralizing antibodies primarily targeting domains EDI and EDII, while EDIII induces potent serotype-specific neutralizing responses.^35^ Antibodies targeting the secreted non-structural protein 1 (NS1), although not directly neutralizing, may reduce disease severity, yet the long-term stability of antibodies against these proteins remains poorly understood.^36^

Flavivirus immunity is influenced by IgG subclass composition (IgG1–4), which differ in their FcγR-mediated immune functions, with IgG1 and IgG3 being pro-inflammatory, IgG2 arising in response to polysaccharide antigens, and IgG4 being immunoregulatory.^37,38^ Minor serum isotypes, including IgM and IgA, also affect immunity, with IgM facilitating early neutralization and complement activation and IgA potentially modulating disease severity.^39–42^ However, limited information exists regarding the long-term persistence and protective roles of these lower-abundance antibodies.

In this study, we characterized the longitudinal dynamics of antibody responses following primary and secondary DENV infections by analyzing antibody isotypes, IgG subclasses, viral antigen specificity, and cross-reactivity profiles. Utilizing multiplex bead-based immunoassays on longitudinal serum samples (<1, 3, 6, and 18 months post-symptom onset) from pediatric dengue cases enrolled in a hospital-based study in Managua, Nicaragua, we investigated the kinetics and durability of these antibody populations. Our findings provide a comprehensive description of antibody subpopulations and their distinct early and long-term kinetic patterns, offering new insights into dengue immunity relevant for vaccine development.

## Results

### Characteristics of participants

In this study, we analyzed plasma samples from 79 pediatric patients with either DENV1 or DENV3 infection. These participants were selected as a convenience sample from a hospital-based observational study in Managua, Nicaragua. We compared the plasma antibody profile of post-primary (1°; n=40) versus post-secondary (2°; n=39) dengue groups. Longitudinal sampling was conducted at 2-4 weeks post-symptom onset (convalescence; Cv), as well as 3, 6, and 18 months (M) post-symptom onset (Supplementary Table S1).

### Distinct antibody kinetics to homologous E protein following primary and secondary DENV infection

To understand the generation and durability of the humoral immune response upon primary versus secondary dengue, we first measured antibodies against the E protein of the infecting serotype (i.e., homologous). The magnitude of each antibody subset was measured as mean fluorescence intensity (MFI) and analyzed after Log_2_ transformation and background (BG) subtraction. A linear model with three distinct temporal periods (Cv-3M, 3-6M, and 6-18M) was applied to each antibody type, and half-life was inferred from the slope of each segment (Supplementary Fig S1, S2). We found that the initial magnitude of homologous E-reactive IgG (E-IgG) at Cv was higher post-2° than post-1° but decreased to a similar level by 18M (Fig 1A). The kinetics of initiating the E-IgG response differed in the Cv-3M period: rising post-1° infection (Cv-3M t_1/2_= −0.34 years, 95% CI=-0.20, −1.11) and waning post-2° infection (Cv-3M t_1/2_= 0.51 years, 95% CI=0.27, 4.37; Fig 1A, B). This trend is driven by the IgG1 and IgG2 subclasses (Fig 1D, E). In contrast, the initial magnitude of anti-E IgA, IgM, and IgG3 responses were higher post-1° than post-2°, peaking at or before convalescence (Fig 1A, B, D, E). Accordingly, the initial waning kinetic (Cv-3M) for these subsets is steeper post-1° than post-2° infection (Fig B, E). The initial levels and kinetics of E-IgG2 and E-IgM compartments demonstrate the greatest differences in 1° versus 2° immunity (Fig 1A, B, D, E).

**Figure 1.**
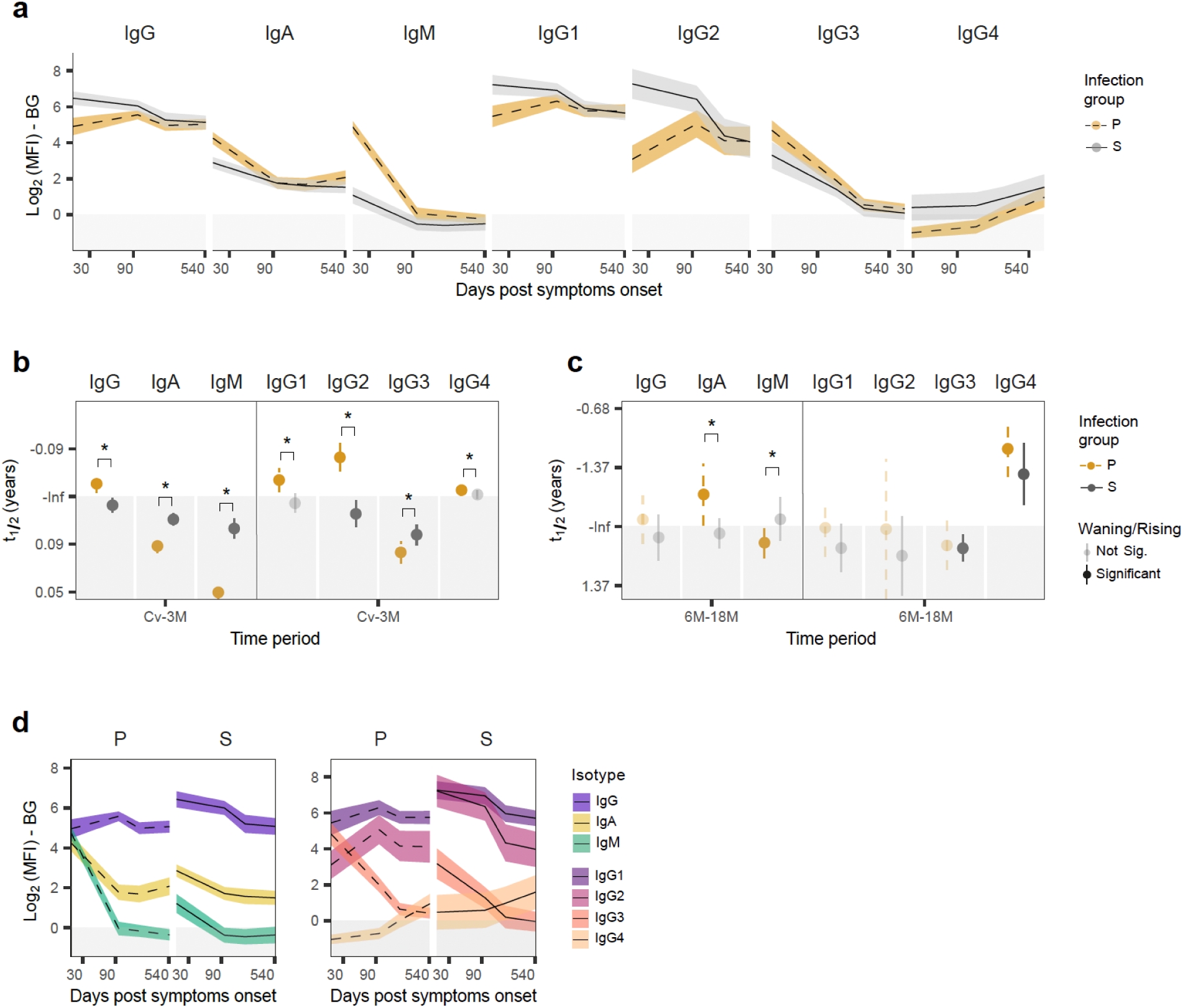
Antibody responses against homologous E protein after primary and secondary dengue. E-binding plasma antibodies against the homologous infecting serotype were measured as Log_2_ of mean fluorescence intensity (MFI) after background subtraction (BG; shaded area). A linear mixed-effects model was applied to estimate a linear rate of decay for three time-period segments: 30-90, >90-180, and >180-540 days post-symptom onset, corresponding to convalescent to 3-month (Cv-3M), 3-6 months (3-6M), and 6-18 months (6-18M) post-symptom onset. **a.** Homologous E-binding antibody levels over time of primary (P; dotted line, yellow) versus secondary (S; solid line, grey) dengue cases with 95% confidence interval for each isotype and IgG subclass. **b.** Short-term antibody half-lives (t_1/2_) estimated from Cv-3M antibody decay rates, and **(c)** long-term t_1/2_ estimated from 6-18M antibody decay rates. Transparent colors indicate stable decay rate with non-estimable infinite (-Inf) t_1/2_ (i.e., Not significant). Solid colors are significantly rising (-) or waning (+; shaded area) antibody kinetics. Significant differences in primary versus secondary decay rates are shown with * for p<0.05. **d.** Comparison of the levels of homologous E-binding antibodies by isotype and IgG subclass after primary and secondary dengue.

We then evaluated antibody durability from 6 to 18M. E-IgG was stable in both 1° and 2° infections, as supported by the IgG1 and IgG2 compartments (Fig 1C, F). Surprisingly, E-IgA increased after 1° (6-18M t_1/2_= −2.52 years, 95% CI=-1.29, −54.82) but not after 2° infections (6-18M t_1/2_= 11.78 years, 95% CI=3.76, −10.37; Fig 1C). Intriguingly, E-IgG4 demonstrated similar kinetics after both 1° and 2° infections and continued to rise long-term both for 1° (6-18M t_1/2_= −1.04 years, 95% CI=-0.77, −1.62; Fig 1D, F) and 2° infections (6-18M t_1/2_= −1.54 years, 95% CI=-0.97, −3.75; Fig 1D, F). Also, we found that by 18M, the magnitude of all anti-E binding antibody subsets was similar across both post-1° and post-2° dengue groups (Fig 1A, D).

Although the anti-E antibody types generally follow the compositional hierarchy observed in healthy human blood (IgG>IgA>IgM, and IgG1>IgG2>IgG3>IgG4), there are some exceptions.^43^ In the Cv-3M phase of the 1° immune response, IgM, IgG1 and IgG3 levels were equivalent (Fig 1D). In the Cv-3M phase of the 2° immune response, IgG2 and IgG1 levels were similar (Fig 1D). In the durable post-2° IgG compartment, E-IgG4 exceeded E-IgG3 levels (Fig 1D).

### Antibody responses against homologous NS1 demonstrate long-term waning

We next assessed how NS1-reactive antibody subsets differ from anti-E antibodies in post-1° versus post-2° groups by measuring the magnitude of antibodies against homologous NS1 (Fig 2). Many aspects were comparable, such as higher levels of NS1-IgG in early 2° compared to 1° responses, a delayed peak of NS1-IgG in 1° compared to 2° responses, and a similar directionality of NS1-IgA and NS1-IgM in both 1° and 2° responses (Fig 2 A, B, D, E). Yet, some distinct patterns emerged. The initial (Cv-3M) rise of NS1-IgG in 1° infections was steeper than the Cv-3M rise in post-1° E-IgG (Fig 2A, D). In 2° infections, NS1-IgG and its IgG1 and IgG2 subclasses demonstrated a sustained peak response through 3M, whereas anti-E IgG declined in this period (Fig 2A, D). Most different was NS1-IgG3, which rose in the 1° Cv-3M period and was stable in 2° Cv-3M, whereas E-IgG3 waned (Fig 2D, E), suggesting a later initiation of NS1 responses than E responses. In the 6-18M period, anti-NS1 IgG, IgG1, IgG2, and IgG3 waned substantially, whereas the anti-E compartments were largely stable (Fig 2C, F). Unlike with E-IgA in the 6-18M phase, NS1-IgA antibodies were stable post-1° and waned substantially post-2° (Fig 2C). At 18M, post-2° anti-NS1 IgG2 and IgG4 levels were higher than post-1° levels (Fig 2D). Thus, long-lasting NS1-IgG wanes, unlike E-IgG, and NS1-IgA demonstrates distinct trends in 1° versus 2° immunity.

**Figure 2.**
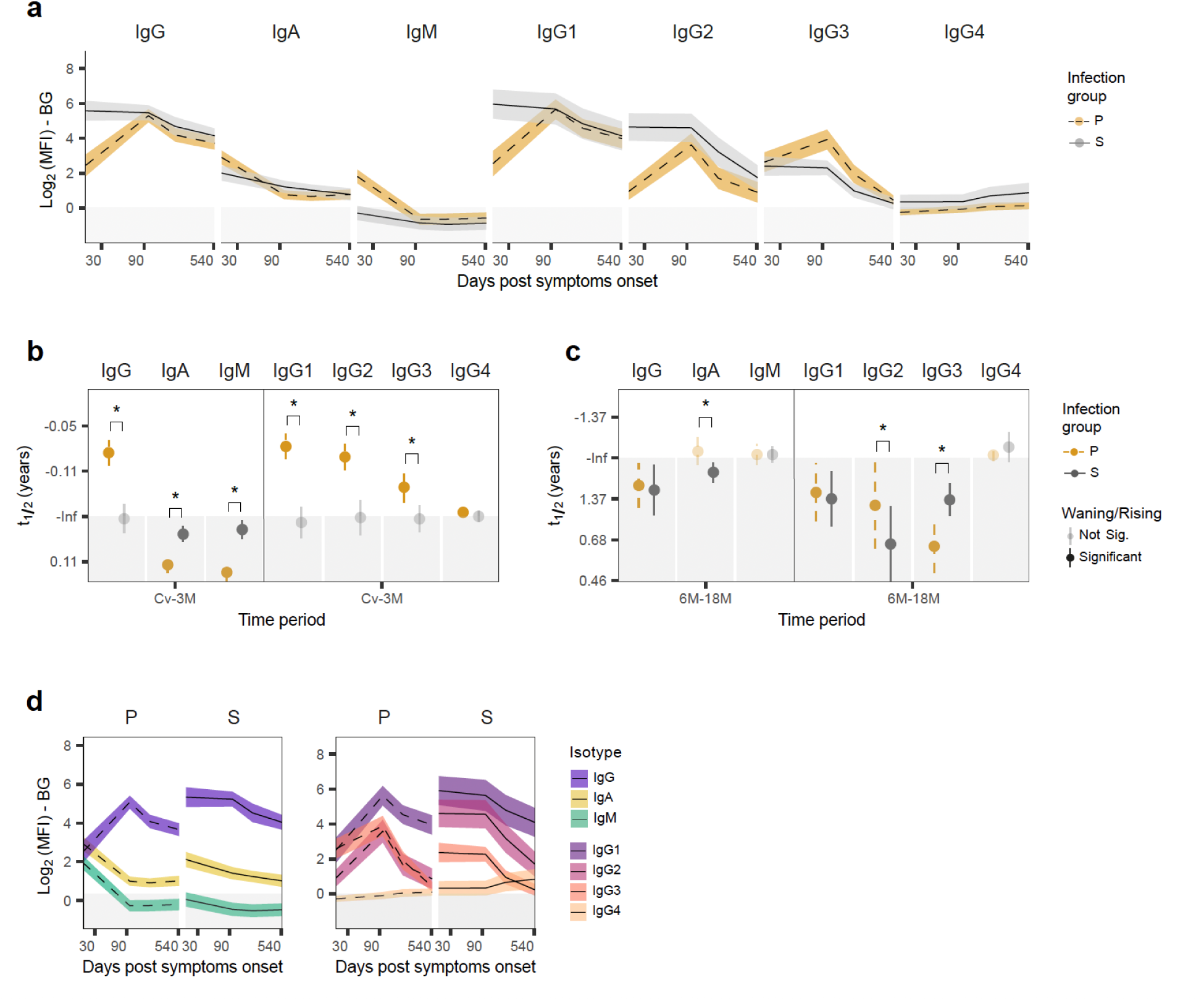
Dynamics of the antibody response against homologous DENV NS1 protein up to 18 months post-symptom onset. NS1-binding plasma antibodies against the homologous infecting serotype (DENV1 or DENV3) were measured as in Figure 1. **a.** Homologous DENV NS1-binding antibody levels over time of primary (P; dotted line, yellow) versus secondary (S; solid line, grey) dengue cases with 95% confidence interval for each isotype and IgG subclass. **b.** Short-term antibody half-lives (t_1/2_) estimated from Cv-3M antibody decay rates, and **(c)** long-term t_1/2_ estimated from 6-18M antibody decay rates. Transparent colors indicate stable decay rate with non-estimable infinite (-Inf) t_1/2_ (i.e., Not significant). Solid colors are significantly rising (-) or waning (+; shaded area) antibody kinetics. Significant differences in primary versus secondary decay rates are shown with * for p<0.05. **d.** Comparison of the levels of homologous NS1-binding antibodies by isotype and IgG subclass after primary and secondary dengue.

### DENV serocomplex E-binding XR antibodies rise post-primary dengue

Since DENV E-binding XR antibodies play a critical role in enhancement or protection in subsequent dengue, we evaluated if this subset demonstrates distinct kinetics. We tested the differences in DENV XR antibody kinetics in post-1° versus post-2° dengue groups. To ensure measurement of XR antibodies only, we analyzed antibodies reactivity to DENV4-E, which is the only DENV serotype to which these patients were unlikely to have been exposed to given DENV epidemiology in Nicaragua during the study period (Supplementary Fig S3, S4). While the early (Cv-3M) kinetics of XR E-IgG were comparable to homotypic E-IgG, the divergence in magnitude of 1° versus 2° IgG responses was more pronounced, particularly in the XR E-IgG1 and E-IgG2 compartment (Fig 3A, B, D, E). Post-2° XR anti-E IgG, IgG1, IgG2, and IgG4 started higher and remained higher (through 18M) than post-1° responses, supporting more efficient selection of a XR antibody repertoire in 2° infections (Fig 3A, D). While XR IgA and IgM anti-E antibodies were higher post-1° than post-2° at convalescence, this may be due to XR epitopes being occupied by higher affinity IgG in 2° infections (Fig 3A).

**Figure 3.**
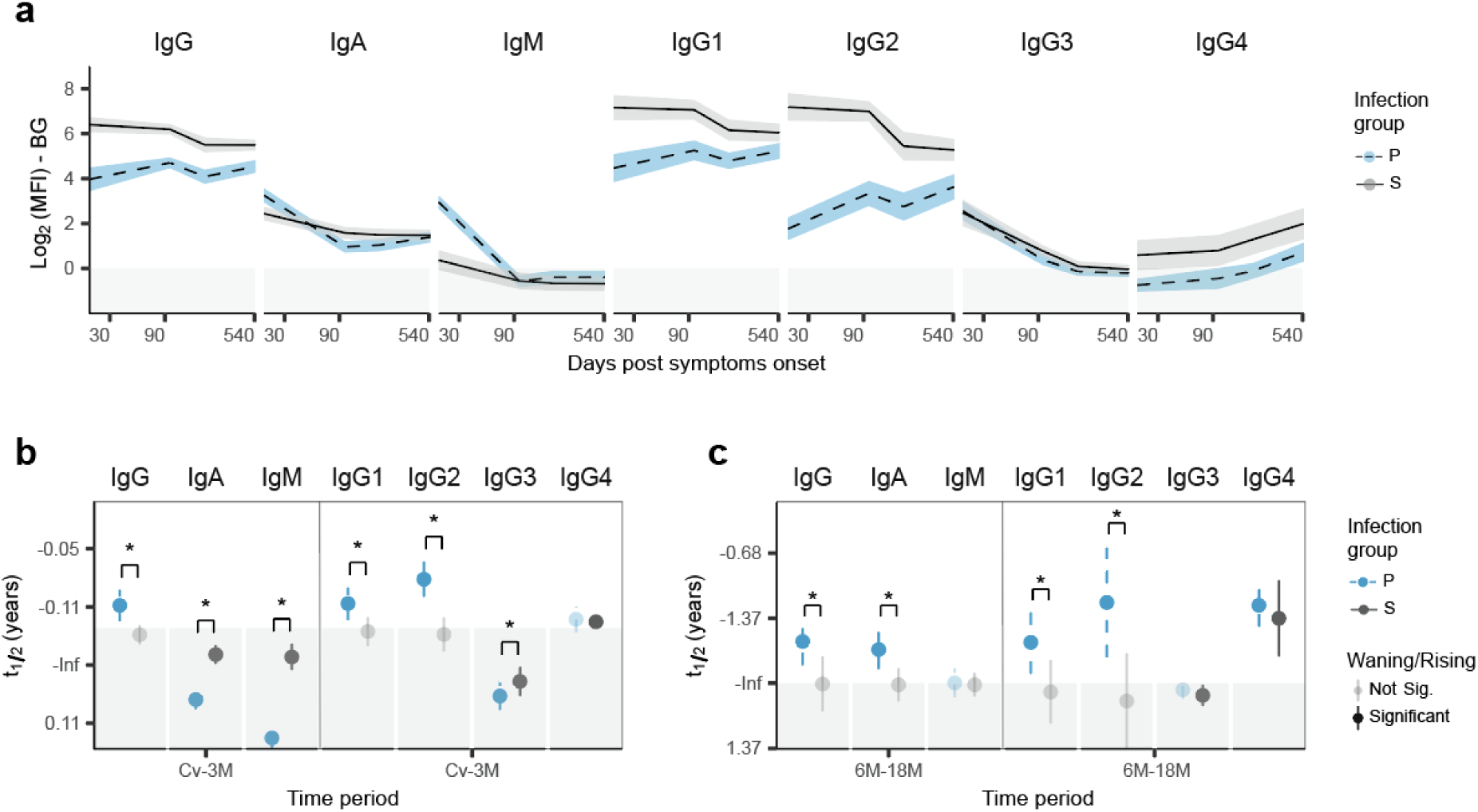
DENV serocomplex cross-reactive anti-E antibodies rise after primary dengue but are stable post-secondary dengue. Cross-reactive (XR) E-binding plasma antibodies were assessed as in Figure 1, against the DENV4 serotype, which was infrequent in our cohort during the time-period of infections sampled here. **a.** XR E-binding antibody levels over time of primary (P; dotted line, blue) versus secondary (S; solid line, grey) dengue cases with 95% confidence interval for each isotype and IgG subclass. **b.** Short-term antibody half-lives (t_1/2_) estimated from Cv-3M antibody decay rates, and **(c)** long-term t_1/2_ estimated from 6-18M antibody decay rates. Transparent colors indicate stable decay rate with non-estimable infinite (-Inf) t_1/2_ (i.e., Not significant). Solid colors are significantly rising (-) or waning (+; shaded area) antibody kinetics. Significant differences in primary versus secondary decay rates shown with * for p<0.05.

The most notable differences between antibody responses to homotypic versus DENV4-XR E were observed in the post-1° 6-18M phase. IgG, IgG1, and IgG2 against XR E rose significantly after 1° infection (6-18M t_1/2_= −2.13 years, 95% CI=-1.36, −4.91), whereas antibodies against homotypic E remained stable and did not rise (Fig 3C, F, Supplementary Fig S5A). Also, after 2° infections, IgG, IgG1, and IgG2 against cross-reactive and homotypic E were both stable (t_1/2_= 140.35 years, 95% CI=-3.36, 3.21; Fig 3C, F). Thus, increasing XR IgG after 1° infection contrasts with stable XR IgG post 2° infection. No waning was observed.

### Low and waning NS1 XR antibody responses

Next, we examined XR NS1 responses, as these antibodies may also contribute to subsequent protection or pathology. As with XR E antibodies, we quantified XR NS1 antibodies using DENV4-NS1, comparing primary and secondary DENV infection groups. However, as cross-reactivity to DENV4-NS1 was often minimal or even undetectable following primary infection, cross-reactivity to DENV2-NS1 was also assessed for this group (Supplementary Fig S6). We found that the level of XR NS1 antibodies was much lower than the total response to NS1 in both 1° and 2° infection groups, indicating that only a small portion of the anti-NS1 response was XR (Supplementary Fig S6A, D). In the 1° infection group at the 6-18M phase, homotypic NS1-IgG waned, but XR NS1-IgG was stable though low (Fig 2, Supplementary Fig S6A, C). In the 2° infection group at the Cv-3M phase, the homotypic NS1-IgG response was stable, whereas the XR NS1-IgG response waned (Fig 2, Supplementary Fig S6A, B, D). At 6-18M phase after 2° dengue, both homotypic and XR NS1-IgG and NS1-IgA waned significantly. Thus, total versus XR NS1-IgG responses demonstrate distinct antibody kinetics.

### Different antibody dynamics by viral antigen and subdomain of the E protein

We next compared total IgG responses by viral antigen to directly test how initiation and durability of antibody kinetics differ for each viral protein. Specifically, we compared NS1, E, and EDIII viral proteins because NS1 is the main non-structural protein, E is the main structural protein, and EDIII is a domain within E that acts as a receptor-binding domain during viral entry into host cells and is the target of strongly neutralizing antibodies (Supplementary Fig S7, S8). While homotypic EDIII and E IgG rose similarly in early (Cv-3M) post-1° infections, XR EDIII-IgG rose significantly more steeply than E-IgG in the same post-1° period, suggesting delayed initiation of XR EDIII-IgG relative to XR E-IgG (Fig 4A, B, C, D, Supplementary Fig S7, S8). In post-2° infections, homotypic and DENV4 XR EDIII-IgG waned significantly more than E-IgG in the Cv-3M period (Fig 4B, D). In the long-term dynamic measured from 6 to 18M, DENV4 XR E-IgG was found to rise significantly in post-1° infection, while EDIII-IgG did not rise (Fig 4E, Supplementary Fig S7, S8), suggesting that the peculiar post-1° rise of IgG antibodies is attributable to E domains I and II. Thus, we found that IgG targeting domain III of the E protein have distinct initiation and post-1° durable kinetics from the overall E protein.

**Figure 4.**
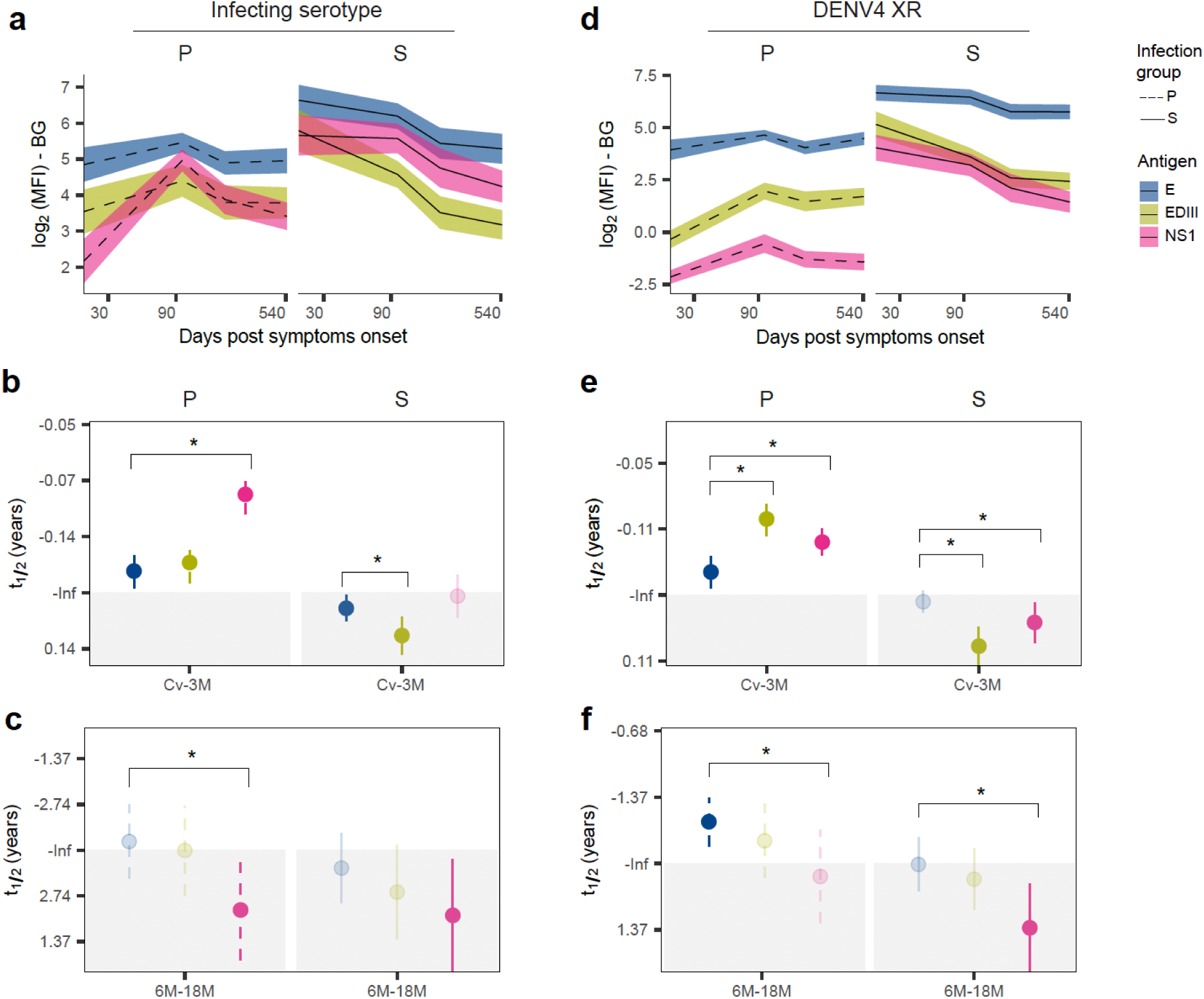
Distinct dynamics of antibodies against E, EDII, and NS1 proteins. DENV-binding plasma IgG levels were measured as in Figure 1. **a.** Comparison of homologous E (blue), EDIII (lime green), and NS1 (pink) binding IgG levels over time in the primary (P; dotted line) versus secondary (S; solid line) dengue groups. **b.** Short-term antibody half-lives (t_1/2_) estimated from Cv-3M antibody decay rates, and **(c)** long-term t_1/2_ estimated from 6-18M antibody decay rates. Transparent colors indicate stable decay rate with non-estimable infinite (-Inf) t_1/2_ (i.e., Not significant). Solid colors are significantly rising (-) or waning (+; shaded area) antibody kinetics. Significant differences in viral antigen-specific decay rates are shown with * for p<0.05. **d.** Comparison of DENV4 cross-reactive (DENV4 XR) E (blue), EDIII (lime green), and NS1 (pink) binding IgG levels over time in the primary (P; dotted line) versus secondary (S; solid line) dengue groups, as well as differences in their short-term **(e)** and long-term **(f)** half-lives.

Next, we compared the dynamics of IgG responses against non-structural (NS1) versus structural (E and EDIII) proteins. We found that post-1° infection, NS1-IgG rose more sharply than E- and EDIII-reactive IgG in the early (Cv-3M) phase, suggesting a delayed initiation of NS1 responses relative to E protein beyond the acute phase (Fig 4A, B). In the long-lasting (6-18M) post-1° IgG compartment, homotypic NS1-IgG waned significantly (6-18M t_1/2_= 2.1 years, 95% CI=1.13, 14.76), whereas DENV4 XR NS1-IgG remained low and stable (Fig 4C, E). This is unlike post-1° DENV4 XR IgG against E protein, which rose significantly 6-18M (Fig 4E). Also, long-lasting post-2° XR NS1-IgG waned significantly (6-18M t_1/2_= 1.41 years, 95% CI=0.84, 4.48) in contrast to the XR E and EDIII IgG, which remained stable (Fig 4C, F). Thus, NS1 and E protein IgG showed significantly different dynamics.

### Distinct kinetics of total versus flavivirus-cross-reactive IgG subsets post-primary dengue

Given that XR antibodies from a 1° immune response pose a risk for development of severe secondary dengue, we directly compared differences in the post-1° response against the infecting serotype (i.e., total) versus XR subsets (Supplementary Fig S3). We defined three XR subsets: two within the DENV1-4 serocomplex (DENV2-XR and DENV4-XR) and another more antigenically distant, outside the DENV1-4 serocomplex (ZIKV-XR; Figure 5A, Supplementary Fig S9). Generally, the magnitude of the IgG response against the infecting serotype was higher than the XR IgG subsets against DENV4 or ZIKV for E, EDIII, and NS1, although the response to E showed the least bias between the infecting versus XR subsets compared to EDIII and NS1 (Fig 5A). Also, the early (Cv-3M) initiation kinetic of the E-IgG response was not significantly different in total versus XR subsets (Fig 5B), whereas the XR EDIII-IgG subsets rose more sharply than the total response, suggesting a delayed establishment of XR EDIII IgG (Fig 5A). Importantly, the 1° immune response was against DENV1 or DENV3 infections. Within the DENV1–4 serocomplex, DENV2 is the most antigenically related to these infecting serotypes, while DENV4 is the most distant. ZIKV, as part of another serocomplex, is antigenically the most divergent among the XR targets analyzed. In alignment with these known antigenic relationships, we observed stepwise decreases in the magnitude of the antibody responses with increasing antigenic distance for EDIII and NS1, but not E (Fig 5). Taken together, these data support that E protein, with the exception of domain III, elicits robust XR antibody responses.

**Figure 5.**
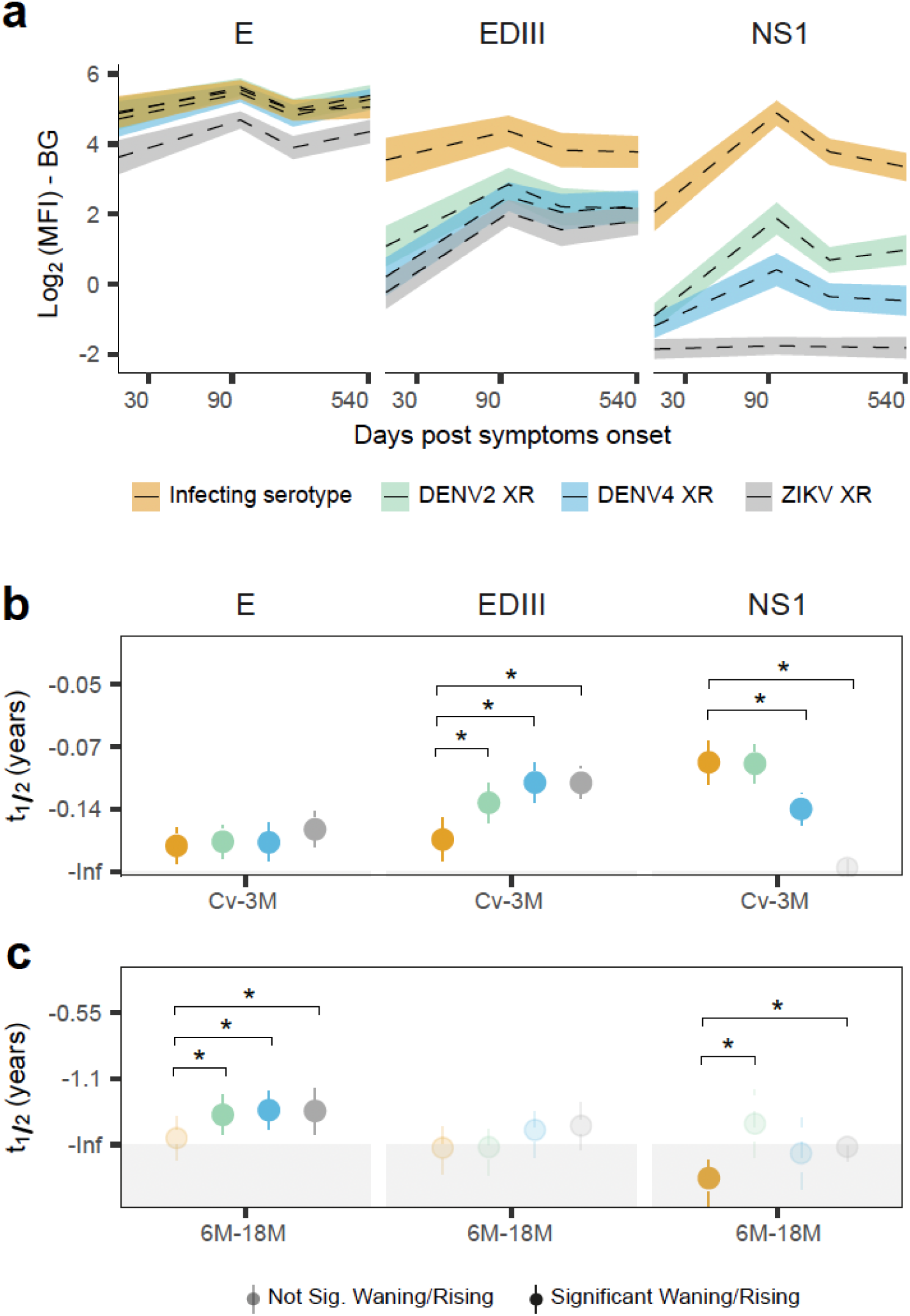
Antigenic hierarchy in the levels and kinetics of cross-reactive antibody responses post-primary dengue. The magnitude of flavivirus E, EDIII, and NS1-binding IgG were measured as in Figure 1. **a.** In the primary dengue group (dotted line), total DENV-reactive IgG against the infecting serotype of DENV1 or DENV3 (yellow), were compared to the magnitude of DENV2 serotype cross-reactive (DENV2 XR; green), DENV4 serotype cross-reactive (DENV4 XR; blue), and ZIKV cross-reactive (ZIKV XR; grey) antibodies over time. For each viral antigen, the short-term antibody half-lives (t_1/2_) were estimated from Cv-3M antibody decay rates **(b)**, and long-term t_1/2_ were estimated from 6-18M antibody decay rates **(c)**. Transparent colors indicate stable decay rate with non-estimable infinite (-Inf) t_1/2_ (i.e., Not significant). Solid colors are significantly rising (-) or waning (+; shaded area) antibody kinetics. Significant differences in cross-reactive antibody subset specific decay rates are shown with * for p<0.05.

Unexpectedly, all long-term (6-18M) post-1° XR E-IgG subsets rose significantly, while the total E-IgG against the infecting serotype remained stable (Fig 5B). In contrast to E-IgG, the long-lasting (6-18M) NS1-IgG against the infecting serotype waned significantly while the XR NS1-IgG subsets were low and stable (Fig 5B). Thus, DENV XR versus homotypic IgG subsets showed distinct kinetics, with XR responses rising (E) or stable (NS1) over time.

### Seropositivity at 18 months post-infection reveals isotype- and antigen-specific patterns

Since seropositivity impacts protection on the population level, we examined seropositivity at 18M for each viral antigen and antibody type after 1° versus 2° infections. Seropositivity was defined as the portion of each 1° or 2° infection group with an antibody response higher than the level of background. Surprisingly, even though IgA and IgM are considered short-lived, we found seroprevalence of IgA and IgM antibodies against the E protein of the infecting serotype post-1° (IgA=100%; IgM=50%) and post-2° was high (IgA=97%; IgM=28%; Fig 6). IgG, IgG1, IgG2, and IgA exhibited the highest seropositivity rates (>50%) for multiple specificities, but with greater seropositivity against viral structural proteins (E and EDIII) than NS1 (Fig 6). Also, IgA, IgG2, and IgG3 demonstrated higher seropositivity for E than for the EDIII subdomain (Fig 6, Supplementary Fig S5B). Moreover, seropositivity declined with greater antigenic distance from the infecting DENV serotype for many antibody subsets (NS1-IgG, NS1-IgA, EDIII-IgG2, E-IgG3), with highest responses towards the infecting serotype, followed by DENV4-XR, and then ZIKV-XR (Fig 6). Taken together, these data indicate distinct antigenic biases by antibody type.

**Figure 6.**
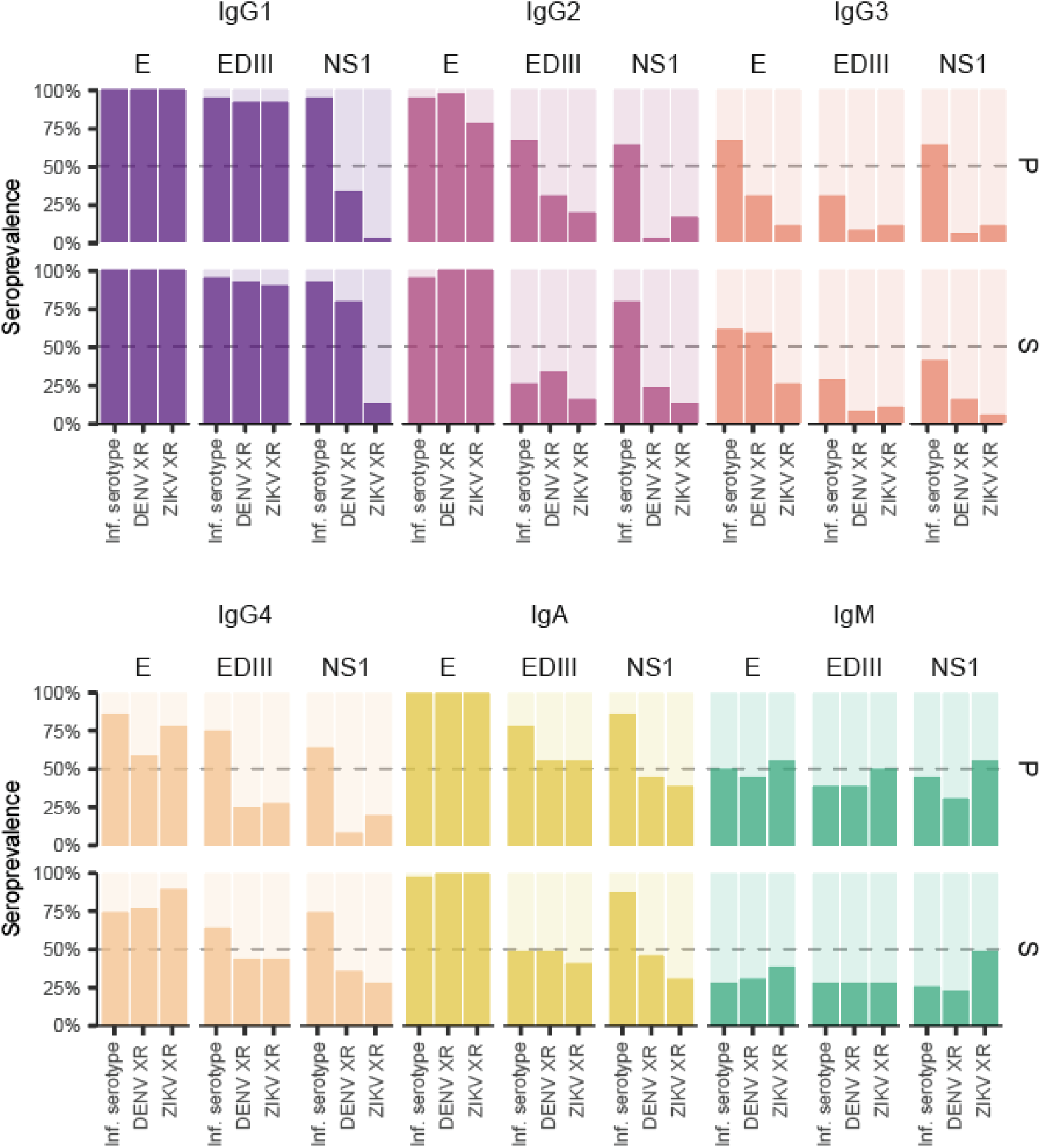
Seropositivity of antibody subsets at 18 months in post-primary versus post-secondary dengue groups. The proportion of the post-primary (P) and post-secondary (S) dengue groups with antibody levels above the level of background was defined as percent seroprevalence for each group. In both groups, antibody subsets are shown by isotypes and IgG subclasses (colors) and stratified by viral antigen (E, EDIII, and NS1) and virus specificity (infecting serotype [Inf. Serotype], DENV4 cross-reactive [DENV XR], and ZIKV cross-reactive [ZIKV XR]). Dotted line marks 50% seroprevalence.

We then assessed which antibody subsets accumulate with exposure. Notably, seropositivity of XR responses was higher in 2° compared to 1° infections (Fig 6). As expected, some antibody types (NS1-IgG1, E-IgG2, E-IgG3, and XR IgG4) showed higher seropositivity in 2° than 1° infection groups (Fig 6, Supplementary Fig S5B). Total IgG, IgG1, and E-IgA did not differ in seropositivity, as they were maximal post-1° response, and IgG3 was low both post-1° and post-2° infections (Fig 6). Other antibody subsets, including all IgM subsets, EDIII-IgA, and EDIII-IgG2 responses, were lower post-2° than post-1° infections, likely due to class-switching and distinct selection of 2° versus 1° immunity in these compartments (Fig 6). In general, the seropositivity of XR responses accumulates with exposure, whereas EDIII seropositivity declines with exposure.

### Inter- and intra-individual variability in DENV antibody responses

Given that intra- and inter-individual variability in antibody responses shape protection at a population level, we sought to examine this variability. With regards to the 18-month DENV4 XR anti-E antibody levels, we found that IgG1 and IgG2 were the most variable across individuals, followed by IgG4 (Figure 7A). Interestingly, IgG1 and IgG2 demonstrated more inter-individual variability after a primary than secondary DENV infection (Figure 7A). This is compatible with a bigger distribution of individual antibody set points after primary than secondary dengue. Next, we examined inter-individual differences in DENV4 XR anti-E antibody dynamics from 6 to 18 months post infection and found that IgG2 kinetics were the most variable across individuals (Figure 7B). Another aspect of inter-individual variation is the directionality of changes in antibody level over time, where antibodies may wane for some individuals, but remain stable or rise for others. We found that 50-75% of the post-1° dengue group have XR anti-E antibodies that rise, whereas only 25% of the post-2° group have XR anti-E antibodies that rise (Figure 7C). This reinforces that rising XR antibodies against E are a more common feature of the post-1° than post-2° dengue response. Finally, we inspected intra-individual variability in antibody levels over time. We found that a high early response was significantly correlated with a high lasting antibody level at 18 months post-1° and post-2° for IgG1 across E, EDIII, and NS1 antigens, and E-IgA (Supplementary Fig S10, S11). Anti-E IgG3 and IgG4 demonstrated this relationship more after 2° than 1° DENV infection, whereas E-IgG2 demonstrated this relationship more after 1° than 2° DENV infection (Supplementary Fig S11). Thus, inter-individual variability in dengue antibody responses may impact population immunity.

**Figure 7.**
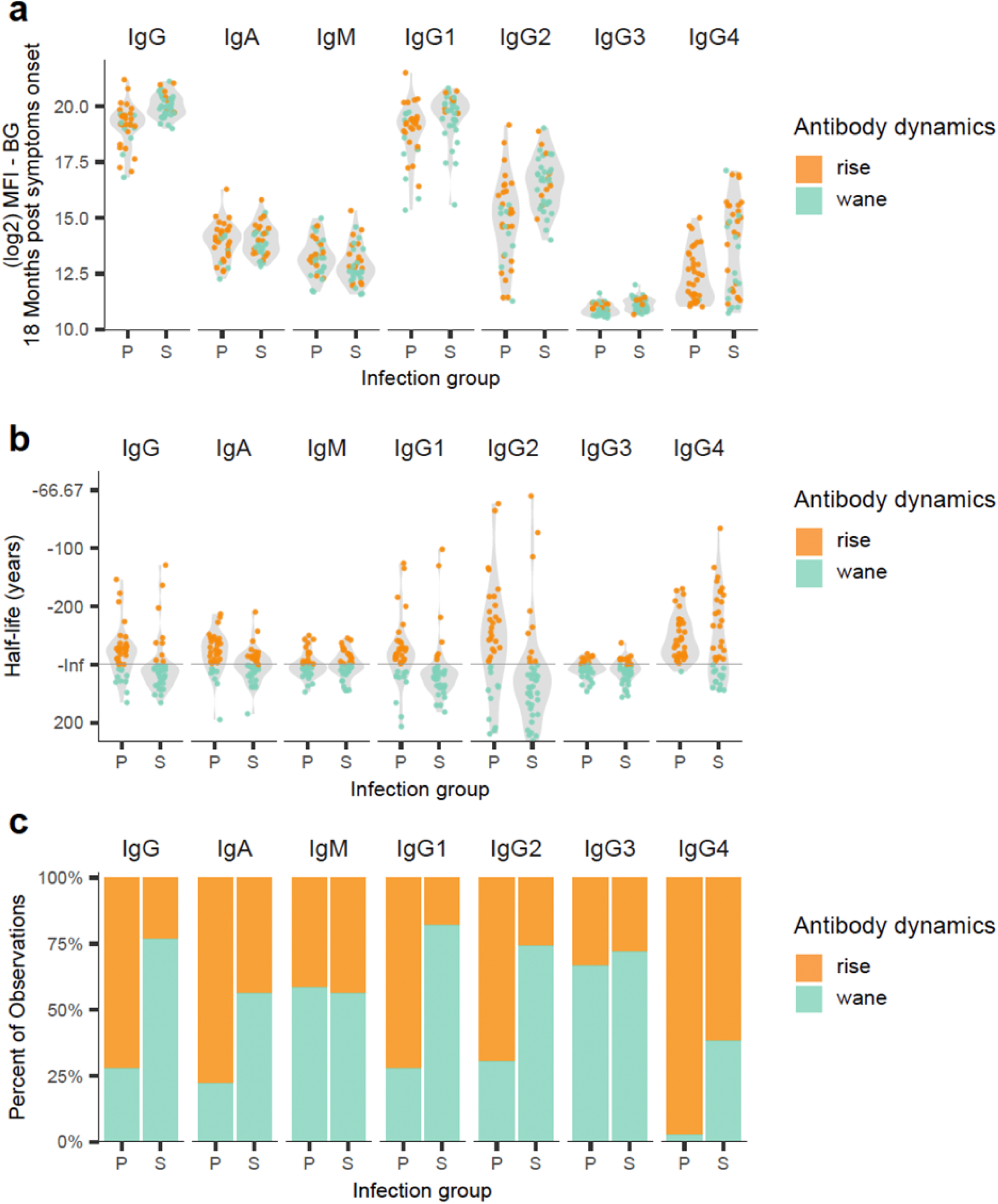
Interindividual variability in DENV4 envelope cross-reactive antibody magnitude and 6-to-18-month post-infection antibody kinetic. DENV4 E-binding plasma antibodies were measured as Log_2_ of mean fluorescence intensity (MFI) after background subtraction (BG; shaded area) in both the post-primary (P) and post-secondary (S) dengue groups. A linear mixed-effects model was applied to estimate a linear rate of decay for 6-18 months (6-18M) post-symptom onset and long-term antibody half-life (t_1/2_) was estimated from the 6-18M antibody decay rates. a. Distribution of the XR anti-E antibody magnitude across individuals at 18 months post infection by infection group. b. Distribution of the XR anti-E antibody t_1/2_ across individuals by infection group. c. Proportion of the post-primary versus post-secondary group with either rising, stable, or waning XR anti-E antibody kinetic from 6-18M for each antibody isotype and IgG subclass. Each dot represents an individual, and color indicates 6-18M antibody kinetics as rising (orange), waning (teal), or stable (green).

## Discussion

In this study, we comprehensively evaluated longitudinal antibody kinetics up to 18 months following 1° and 2° DENV infections, detailing responses for 84 antibody subsets defined by viral antigen, isotype, subclass, and cross-reactivity. This work highlighted several novel conceptual insights into flavivirus immunity. First, it challenges the traditional assumption that cross-reactive antibody levels wane after primary infection. Instead, we observed subsets (IgG1, IgG2, IgA) rise from 6 to 18 months post-infection. Second, it demonstrates that antibody dynamics varies not only by infection history but also by antibody specificity (i.e., infecting serotype versus cross-reactive) and epitope/domain. Furthermore, we detected substantial DENV-binding IgA, IgM and IgG3 seropositivity at 18 months post-infection, indicating longer persistence than expected for these isotypes. Together, these observations suggest that the humoral response to DENV is not homogenous but comprises multiple differently regulated antibody subsets, each with unique kinetic profiles influenced by prior infection history and other intrinsic factors.

We evaluated post-1° XR antibody responses because waning XR IgG is thought to enhance subsequent heterologous DENV infection. Surprisingly, we observed a significant rise in XR E-IgG, particularly the IgG1 and IgG2 subclasses, from 6 to 18 months post-1° DENV infection. This finding contrasts with prior reports, which indicate stabilization or decline in antibody levels.^27,44^ Several factors may explain this discrepancy. First, differences in assays used and in statistical power may have impacted kinetic changes detected in prior studies. Second, aggregation of antibody responses without distinction of DENV homotypic versus XR antibodies might have masked divergent trends. Specifically, we observed stable post-1° long-term total anti-E IgG (i.e., against the infecting serotype) but a rising XR antibody subset; thus, we infer that the type-specific antibody response may wane. This indicates a change in the antigenic composition of post-1° DENV-reactive plasma IgG over time. Interestingly, a similar rise in anti-DENV antibodies has been documented following 1° ZIKV infection.^16,28^ Here we show that this phenomenon also occurs in the reciprocal direction (i.e., DENV and ZIKV XR antibody rise after 1° DENV infection), suggesting that the rising kinetic of XR antibodies may be a broader feature of 1° flavivirus infections. It is important to note that our samples are derived from an endemic region with sustained DENV transmission. Thus, boosting due to subclinical re-exposures likely contributes to the antibody kinetics observed here.^45^ Antibody dynamics following a single primary exposure in travelers or in vaccinees from non-endemic regions may follow different trajectories.

Given that low to intermediate levels of XR IgG induced by 1° flavivirus infections can facilitate DENV ADE, ^8–10^ our data up to 18 months post-infection suggest an alternate perspective on how this may happen due to a low level of rising, rather than waning DENV XR antibodies. Moreover, while XR E-IgG rises post-1° infection, XR EDIII-IgG remains stable. This suggests that the long-term post-1° rise in XR E-IgG is attributable to XR epitopes on E domains I and II, but not domain III. Indeed, EDIII is the least conserved and harbors type-specific epitopes.^33,46,47^ Another factor that might modulate the risk for disease severity is the significant rise of XR E-IgA and XR NS1-IgA antibodies from 6 to 18 months post-1° DENV infection. While E-IgA has previously been associated with protection, we recently linked elevated levels of NS1-IgA with increased severity of dengue disease.^48,49^ Our data indicate a need to monitor XR E-IgG, XR E-IgA, and XR NS1-IgA over years to better understand how this may modulate the potential for antibody-mediated risk.

In contrast to a 1° DENV immune response, post-2° DENV antibodies are thought to be broadly protective against the DENV1-4 serocomplex.^21^ Consistent with earlier findings, our data confirm distinct antibody trajectories across 1° versus 2° DENV infections, notably the earlier and higher peak of anti-E IgG, particularly IgG1 and IgG2, and the sustained higher levels of XR E-IgG (IgG1, IgG2, IgG4) at 18 months after 2° compared to1° DENV infections.^38^ In the post-2° DENV immune response, we and others have found that the difference in the magnitude between E-IgG and EDIII-IgG gets wider over time, unlike a 1° response, suggesting that the 2° immune response more efficiently selects for antibodies targeting epitopes on E outside of EDIII.^44^ This is concordant with prior studies reporting the selection of potently neutralizing quaternary antibodies, which have footprints on E domains I and II.^30,50,51^ Thus, in alignment with many studies, we find that the 2° DENV immune response selects a XR antibody repertoire more efficiently than a 1° response, which may contribute to subsequent protection from severe disease.

Antibodies against NS1 have been minimally studied longitudinally, but NS1-IgG may participate in protection by limiting viral dissemination and vascular leak during subsequent infection. ^36,52,53^ Alternatively, XR NS1-IgA can promote dengue pathogenesis, by stimulating neutrophil activation in secondary infection. We draw several important observations regarding NS1 antibodies. First, NS1-IgG against the infecting serotype wanes significantly after both 1° and 2° DENV infection, unlike E-IgG. Second, the magnitude of the 1° immune response was higher against the infecting serotype than against XR antigens, and this bias is greater for NS1 than for E protein, suggesting that NS1 antibodies are likely predominantly type-specific. Third, the XR NS1-IgG antibodies were stable even though overall NS1-IgG waned, suggesting that that type-specific anti-NS1 antibodies waned over time. Finally, we observed a delayed initiation of antibodies to NS1 relative to E protein, consistent with a prior study.^54^ This delayed onset, along with the differential durability and type-specificity of NS1 antibodies, highlights fundamental differences in the kinetics and composition of antibody responses against structural versus non-structural viral proteins and aligns with findings from HIV, influenza, and SARS-CoV-2.^55–58^

With regard to the early kinetic reflecting the initiation of the immune response, our data suggests a temporal hierarchy in the establishment of humoral immunity. For a given antigen, antibodies binding to the infecting serotype displayed different kinetics than XR antibodies, suggesting that type-specific and XR antibodies follow different dynamics. Further, following 1° infection, XR IgG responses measured against XR EDIII and XR NS1 increased more steeply between convalescence and 3 months post-infection than responses measured using the homologous antigen from the infecting serotype. This suggests a delay in the initiation of the cross-reactive response relative to homologous response. Surprisingly, kinetic differences were observed even among XR antibodies targeting distinct domains within the same antigen. For instance, following 1° infection, EDIII-specific XR antibodies arise later than those targeting EDI/II. During 2° infection, the pattern reversed: EDIII-reactive antibodies waned more rapidly than E-binding antibodies. These different kinetics suggest differential selection of epitope-specific populations over time.

Our approach of deconvoluting the antibody response by related viruses reveals important insights regarding sub-antigen epitopes. It is intriguing that upon primary infection, the kinetics of antibodies targeting a given domain or protein (e.g., EDIII or NS1) differed depending on the virus used as antigen. Although these proteins share homologous structures, they present different epitope landscapes due to amino acid divergence. As a result, antigens from the infecting serotype versus DENV4/2, or even ZIKV capture some overlapping and some non-overlapping subsets of antibody, such that the difference in antibody dynamics reflect epitope-level differences within the cross-reactive antigen.

Altogether, these findings indicate that antibodies targeting different epitopes may exhibit distinct kinetics, and this has important implications for antibody-mediated disease risk. If epitope-specific antibodies have distinct dynamics, it is possible that protective and enhancing antibodies can have different dynamics. Since the protective or enhancing potential of cross-reactive antibodies depends not only on their magnitude but also on their qualitative composition (e.g., the epitopes they recognize), differential kinetics among these subsets may have important implications for disease risk. Our findings suggest that the composition of the cross-reactive antibody pool is dynamic between infections. Thus, the relative balance of protective versus enhancing antibodies could evolve over time, resulting in shifts in disease risk that may not reflect uniform antibody trajectories, but rather the differential persistence or waning of specific epitope-targeting populations.

Finally, comprehensive evaluation of minor human plasma immunoglobulin constituents (e.g., IgA, IgM, IgG3, IgG4) revealed surprising insights. First, we identified long-lasting antiviral IgA, IgM, and IgG3 antibodies, indicated by substantial seropositivity at 18 months post-1° and post-2° DENV infection. While these antibody types are often considered short-lived and a hallmark of recent infection, we find this is not the case when measured with sensitive and specific assays. ^59,60^ Others have reported such long-lasting flavivirus antibodies as well.^61,62^ This is particularly relevant, as long-lasting antibody subsets may contribute to protection versus pathology in subsequent infection.^63^ Previously, we found that long-lasting E-IgM and antibody-dependent complement deposition and virolysis were associated with protection from subsequent symptomatic DENV3 infection and severe dengue disease.^42,64^ Second, we found that E-IgG4 continues to rise from 6 to 18 months after 1° and 2° DENV infection and demonstrated higher seropositivity after 2° than 1° DENV infection. Recent studies indicate that class-switch toward IgG4 regulates the immune response to lessen antibody-dependent phagocytosis and complement deposition, which may aid in minimizing pathology and inflammation in subsequent DENV infection.^65^ In fact, we have consistently observed pre-infection IgG4 antibodies to both E and NS1 to be associated with protection from both symptomatic DENV3 infection and severe dengue disease.^42,64^ Third, IgA, IgG2, IgG3, and IgG4 demonstrated higher seropositivity at 18 months post-infection for E than EDIII or NS1, indicating that antigenic biases exist by antibody type. This may result from differences in how each viral antigen is exposed and processed during infection, as well as from distinct cytokine environments elicited by structurally and spatially distinct antigens (virion versus NS1), which can differentially prime the immune response.

In addition, certain isotype–antigen combinations within XR antibody subsets, such as E-IgG2 and E-IgG4, show marked interindividual variability at 18 months post-1° infection. Also, a subset of individuals remain seropositive to cross-reactive antigens with the presence of minor isotypes like IgA, IgM, IgG3, and IgG4, while others had undetectable levels. It is possible that there may be more IgA and IgM than we observed as some experimental signal may be lost due to the abundance of IgG in human plasma. Such heterogeneity may have important implications for long-term immunity by influencing risk or protection against subsequent heterologous infections. Accordingly, these isotype-specific responses may segregate the population into distinct responder groups and serve as valuable targets for correlates of protection studies, as they are likely to reflect meaningful biological variation.

A significant strength of our study is the detailed characterization of antibody kinetics across multiple dimensions of the DENV immune response in natural infections, including antibody isotype and subclass, antigen specificity, and virus cross-reactivity, using multiplex assays that offer higher resolution than traditional assays. Additionally, our longitudinal sampling approach and statistical modeling provide rigorous estimates of antibody dynamics and durability. However, our study also has limitations, such as a relatively small sample size, which may result in lower statistical power to detect differences in waning over time. Our convenience sampling design may limit generalizability. Infection histories from 2° individuals were unknown, and it is possible that antibody kinetics are further modulated by the exact number of flavivirus exposures or the serotype(s) of prior infection. Also, we evaluated DENV1 and DENV3 serotype infections, and dynamics may differ somewhat for other DENV serotypes. Finally, our analyses focused on antibody binding and did not directly assess functional or neutralizing activity, which may more closely reflect protective immunity. Of note, our longitudinal sampling timepoint is up to 18 months post-infection, and longer-term dynamics that may contribute to protection versus enhancement of subsequent dengue must be confirmed.

In conclusion, this study reveals complex antibody dynamics following 1° and 2° DENV infections. The rise of XR antibodies post-1° infection and the differences in antibody trajectories of E, EDIII, and NS1 add critical insights to our understanding of dengue immunity. These findings are important for vaccine design, diagnosis, epidemic surveillance, and modeling.

## Methods

### Study design

The hospital-based pediatric dengue study is an ongoing longitudinal investigation at the Hospital Infantil Manuel de Jesús Rivera (HIMJR) in Managua, Nicaragua.^45,46^ Since 2005, children aged 0–14 years who present within 1–7 days of symptom onset are invited to participate if they meet case definitions for dengue, Zika, or chikungunya. Following consent or assent, participants undergo standard clinical evaluations and, if needed, hospitalization based on established criteria.^13^

Blood samples are collected at the time of hospital presentation (acute phase), daily for days 1–3 of illness, and again in early convalescence (14–28 days post-onset).^46^ Laboratory testing confirms infections using RT-PCR, virus isolation, and/or serological assays for dengue virus (DENV), Zika virus (ZIKV), and chikungunya virus (CHIKV). Clinical data, comprising around 150 variables (including fluid intake/output), are recorded every 12 hours and digitized via a double-data–entry protocol with daily and weekly quality control checks. A standardized questionnaire captures demographic and epidemiological information, ensuring consistency and completeness. In addition, children may consent to a longitudinal follow-up of convalescent samples at 3, 6, 12, and 18 months after illness onset. These follow-up visits collect healthy plasma and PBMC samples. For this study, samples were collected from August 24, 2010, to November 26, 2013.

Laboratory confirmation of DENV infection specifically involves either seroconversion of DENV-specific IgM antibodies between acute- and convalescent-phase samples with MAC-ELISA, a fourfold or greater rise in total antibodies measured by inhibition ELISA, virus isolation in C6/36 Aedes albopictus cells, or RT-PCR amplification of viral RNA. DENV serotypes (DENV1–4) are identified by virus isolation or RT-PCR.^66–68^ A primary DENV infection is classified by an iELISA titer below 20 in acute-phase samples and/or below 2560 in convalescent-phase samples; conversely, secondary infections are indicated by titers of 20 or higher in acute-phase samples and/or 2560 or higher in convalescent-phase samples.^46^ From 2016 onwards, differential diagnosis from ZIKV infections is performed considering ZIKV RT-PCR, ZIKV IgM and ZIKV inhibition ELISA seroconversion.

This study was reviewed and approved by the Institutional Review Boards of the University of California Berkeley, the Ministry of Health of Nicaragua, and the HIMJR. Parents or legal guardians of all participants provided written informed consent, and participants 6 years of age and older provided assent.

### Multiplex assays

Recombinant envelope (E) and NS1 proteins of DENV1–4 and ZIKV were purchased from the Native Antigen and randomly biotinylated. Envelope domain III (EDIII) site-specifically biotinylated of DENV1–4 and ZIKV were provided by Lakshmanane Premkumar, University of North Carolina, Chapel Hill. Antigens were coupled to avidin-coated MagPlex beads and pooled for use in a multiplex assay. Briefly, diluted plasma samples (four consecutive fourfold dilutions beginning at 1:50) were incubated with the antigen-coupled microspheres in 384-well plates for 90 minutes at 37°C under continuous shaking at 1200 rpm. Following incubation, the plates were washed with PBS containing 0.1% bovine serum albumin and 0.02% Tween 20 using a Tecan Hydrospeed automatic magnetic washer. Antigen-specific Ig were detected by adding phycoerythrin (PE)-conjugated mouse anti-human IgG, IgM, IgA or IgG1-4 as indicated (SouthernBiotech). Median fluorescence intensity (MFI) was measured on an iQue3 (Intellicyt). Five DENV-naïve human serum samples collected during the acute phase of a non-DENV febrile-illness were included on each plate as a negative control. Background-subtracted MFI values were calculated using average of naïve individuals + 3SD. MFI were included when ≥ 50 beads were acquired per region.

### Statistical analyses

To compare primary and secondary infections, as well as all isotypes and antigens at the same dilution, the 1:800 dilution was used. We conducted three primary comparisons—(1) primary versus secondary infection status (IR), (2) viral antigen (E, EDIII, and NS1), and (3) viral response (against incoming serotype, DENV cross-reactive, and ZIKV cross-reactive)—while carefully accounting for the complexity of the data. Each factor was examined simultaneously, considering the other relevant factors by embedding analyses within multiple stratifications (e.g., primary versus secondary infection comparisons were analyzed separately for each antigen, viral response category, and antibody isotype and subclass). Analyses were conducted using linear mixed-effects models with the general form:

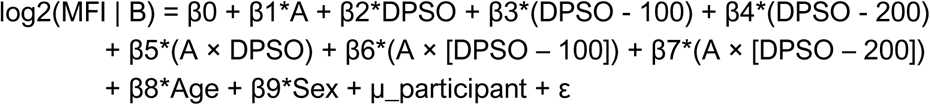

where A is the single primary factor of interest (IR, antigen, or viral response), and B represents the outcome level of antibody as Log_2_(MFI). We incorporated piecewise linear splines for days post-symptom onset (DPSO) at knots of 100 and 200 days to capture immunologically known phases: an initial rapid decay in antibodies, followed by stabilization, and then a long-term homeostasis at later times. Therefore, β2 captures the slope (rate of change in antibody levels) from day 0 to 100 (∼3 months), β3 is the change in slope from 100 to 200 days (3 to 6 months), and β4 is the change in slope from 200 to 540 days (6 to 18 months). A random intercept (µ_participant) was included for each participant to account for repeated measures over time from the same individual. Age and sex were included as covariates in all models. To robustly quantify uncertainty, we performed case bootstrap resampling at the participant level refitting each model 1000 times.^69^ For each bootstrap replicate, marginal predictions of log₂(MFI) were computed at 20 days, 3 months (≈100 days), 6 months (≈200 days), and 18 months (≈550 days), enabling comparisons of antibody magnitude distributions across these time points. Additionally, we derived daily rates of MFI change within each piecewise segment (20– 100 days, 100–200 days, and 200–550 days) and averaged these slopes across replicates, providing a quantitative basis for comparing antibody kinetics (slopes) across groups. These daily rates of change in log₂(MFI) were subsequently transformed into half-lives using the formula:

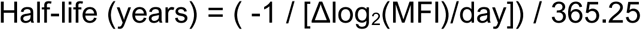

Under this convention, a half-life of zero implies an infinite half-life (no net change in antibody concentration), negative values represent a rising antibody response ("negative half-life"), and positive values reflect waning antibody concentrations ("positive half-life").

Finally, seroprevalence, defined as the proportion of samples exceeding three standard deviations above the naïve mean value, and was calculated at 18 months post-infection for each factor level (infection status, antigen type, viral response category, antibody isotype/subclass). All statistical analyses were conducted in R (version 4.3.1) using the following packages: lme4, lmeresampler, ggeffects, lspline, emmeans, dplyr, and foreach. Confidence intervals (95%) for predictions and slopes were derived using the bootstrap-normal approximation. Code for statistical analyzes can be found at 10.5281/zenodo.16743227.

## Data Availability

All data produced in the present study will be available upon publication of the manuscript and reasonable request to the authors

## Acknowledgements

We are deeply grateful to the participants of the Pediatric Dengue Cohort Study (PDCS) and their families for their continued commitment. We acknowledge the dedication and invaluable contributions of the past and present study team members at the Hospital Infantil Manuel de Jesús Rivera (HIMJR), the Sustainable Sciences Institute in Nicaragua, and the Laboratorio Nacional de Virología at the Centro Nacional de Diagnóstico y Referencia. We thank Dr. Lakshmanane Premkumar for generously providing recombinant antigens and for his technical support.

## Author contributions

S.B. and E.H. conceived the project. S.B. designed and carried out the experiments; S.B. and J.V.Z. performed data analysis; and S.B., T.S., J.V., and E.H. contributed to interpretation of the results and wrote the manuscript. J.V.Z. managed the laboratory databases and organized the data for analysis. A.G., E.D., R.M.H., and J.H. carried out experiments. A.B. directed the cohort study. E.H. obtained funding. All authors provided critical feedback and reviewed the manuscript. This work was supported by grants U19AI118610 (EH, Ana Fernandez-Sesma) and P01AI106695 (EH) from the National Institute for Allergy and Infectious Diseases of the National Institutes of Health (NIAID/NIH). T.S. was supported by the HHMI Hanna H. Gray Fellowship (Grant ID: GT16780). The PHDS was supported by NIAID/NIH grants R01AI099631 (AB) and U54 AI065359 (AB; Program Director Alan Barbour).

## Materials & Correspondence

Please address the correspondence and material requests to Eva Harris.

