## Supplementary material for "Longitudinal antibody profiling after dengue reveals distinct dynamics by antibody specificity over 18 months"

**Supplementary Table 1. Study participant characteristics by post-primary versus post-secondary dengue groups.**

| | | Primary | | Secondary | |
|---|---|---|---|---|---|
| | All | DENV1 | DENV3 | DENV1 | DENV3 |
| Participants | 79 | 20 | 20 | 16 | 23 |
| Age (mean (SD)) | 8.6 | 7.6 | 7.0 | 10.4 | 9.7 |
| Female | 32 | 10 | 10 | 5 | 8 |
| Male | 47 | 10 | 10 | 11 | 15 |
| DHF | 13 | 0 | 0 | 3 | 10 |
| Days from Symptom onset (Convalescent) *(median [IQR])* | | 16.00 *[13.50, 17.00]* | | 16.00 *[15.00, 18.50]* | |
| Days from Symptom onset (3 months) *(median [IQR])* | | 107.50 *[101.00, 114.25]* | | 111.00 *[103.50, 122.00]* | |
| Days from Symptom onset (6 months) *(median [IQR])* | | 188.00 *[186.75, 194.25]* | | 189.00 *[186.00, 196.50]* | |
| Days from Symptom onset (18 months) *(median [IQR])* | | 562.50 *[551.25, 578.75]* | | 557.00 *[553.00, 568.00]* | |

a.

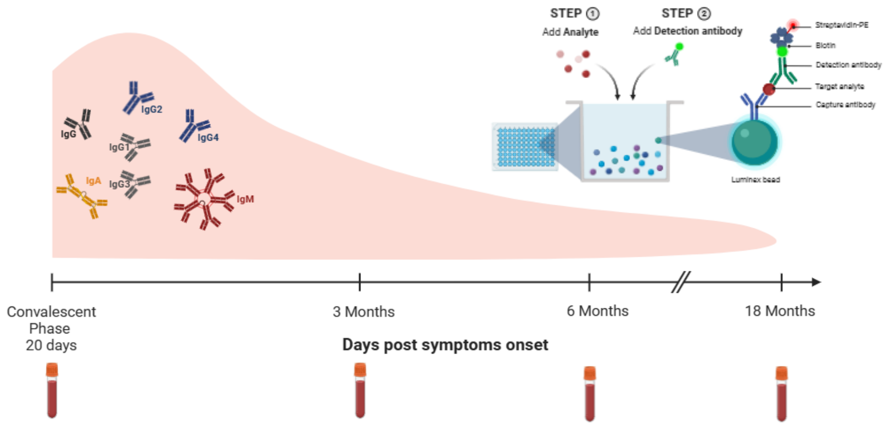


b.

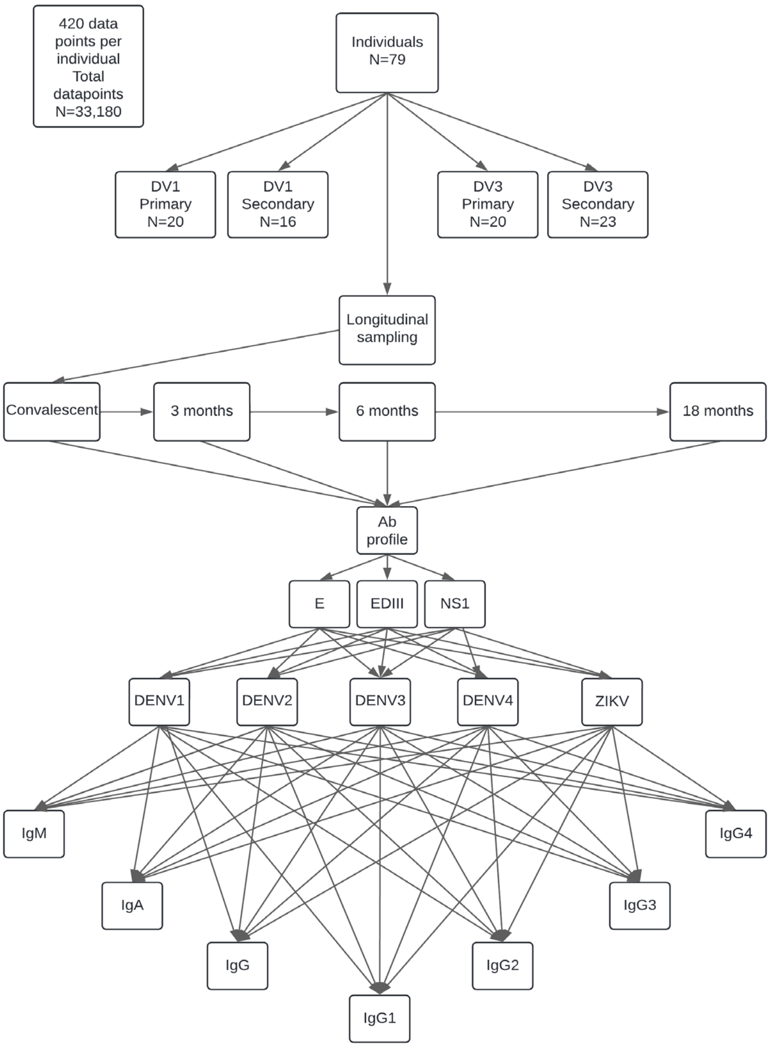


**Supplementary Figure 1. Schematic of experimental and analysis approach. a.** The experimental approach utilized longitudinal plasma samples collected at convalescence, 3 months, 6 months and 18 months post-onset of dengue symptoms. Antibody levels were measured using a multiplexed bead-based assay where the antigens were conjugated to fluorescent beads, co-incubated with plasma, and the isotype or IgG subclass or antigen-bound antibodies was detected with a fluorescent secondary antibody. **b.** In total, 420 data points were collected for each of the 79 individuals, yielding 33,180 data points in total. Given the longitudinal sampling, there are multiple relationships between the key variables, which are indicated with arrows.

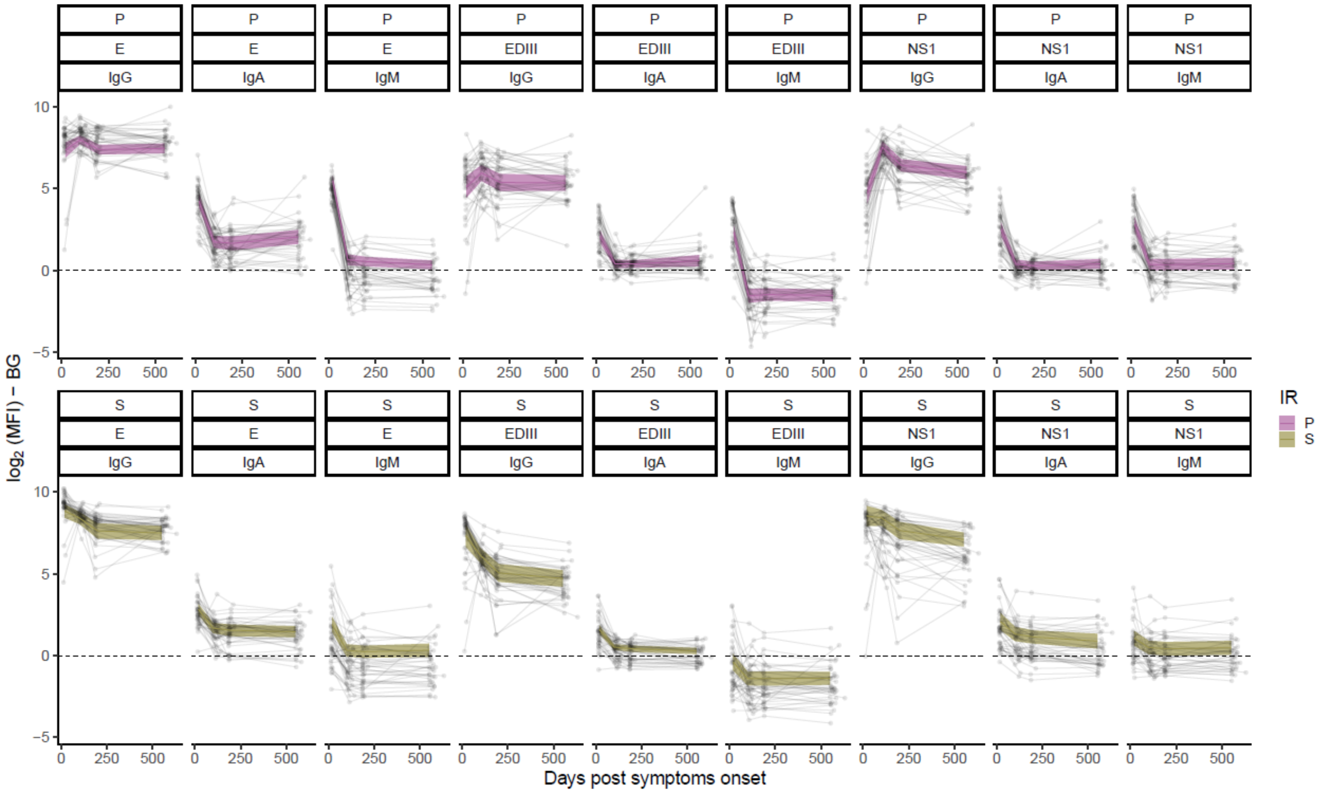


**Supplementary Figure 2. Model fit atop individual antibody trajectories for homologous antibody responses against the infecting serotype.** The magnitude of binding antibodies were measured as $\mathrm{Log}_2$ of mean fluorescence intensity (MFI) after background subtraction (BG; shaded area). A model was applied to estimate a linear rate of decay for three time-period segments: 30-90, >90-180, and >180-540 days post-symptom onset. Raw data of IgG, IgA, and IgM antibody levels against E, EDIII and NS1 over time for each individual with respect to antibody responses against the infecting serotype are shown. Model fit of the primary group is shown in pink (P) and the secondary group in beige (S), with 95% confidence intervals. The dashed line represents the background derived from DENV-naïve plasma samples.

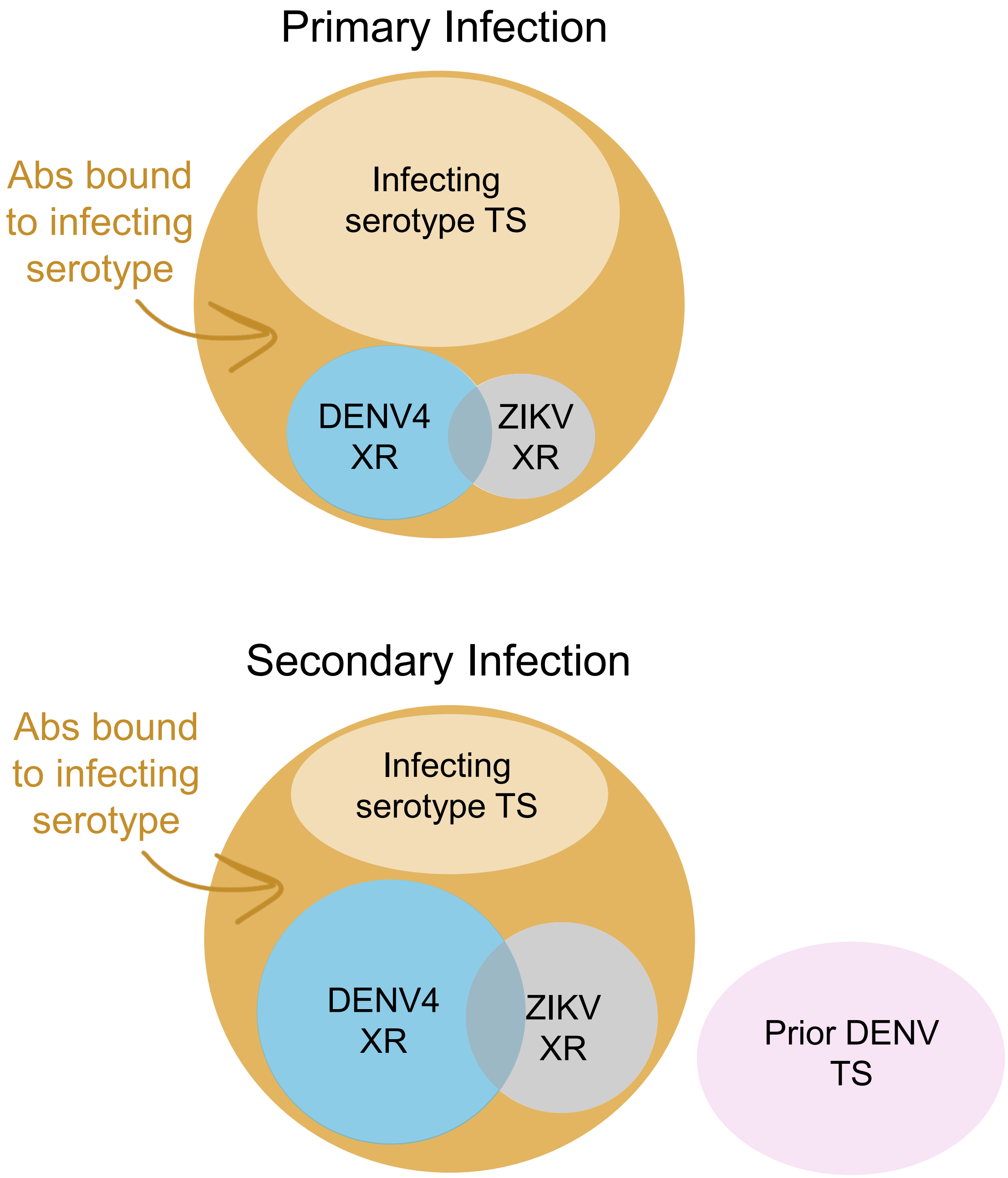


**Supplementary Figure 3. Schematic representation of key concepts regarding cross-reactive antibody sub-populations during a primary versus a secondary immune response.** Notably, antibodies against the infecting serotype (i.e., homologous) do not represent only type-specific (TS) antibodies, which are not possible to measure with our approach. The size of the circles represents the magnitude of each cross-reactive (XR) subset as per our current understanding.

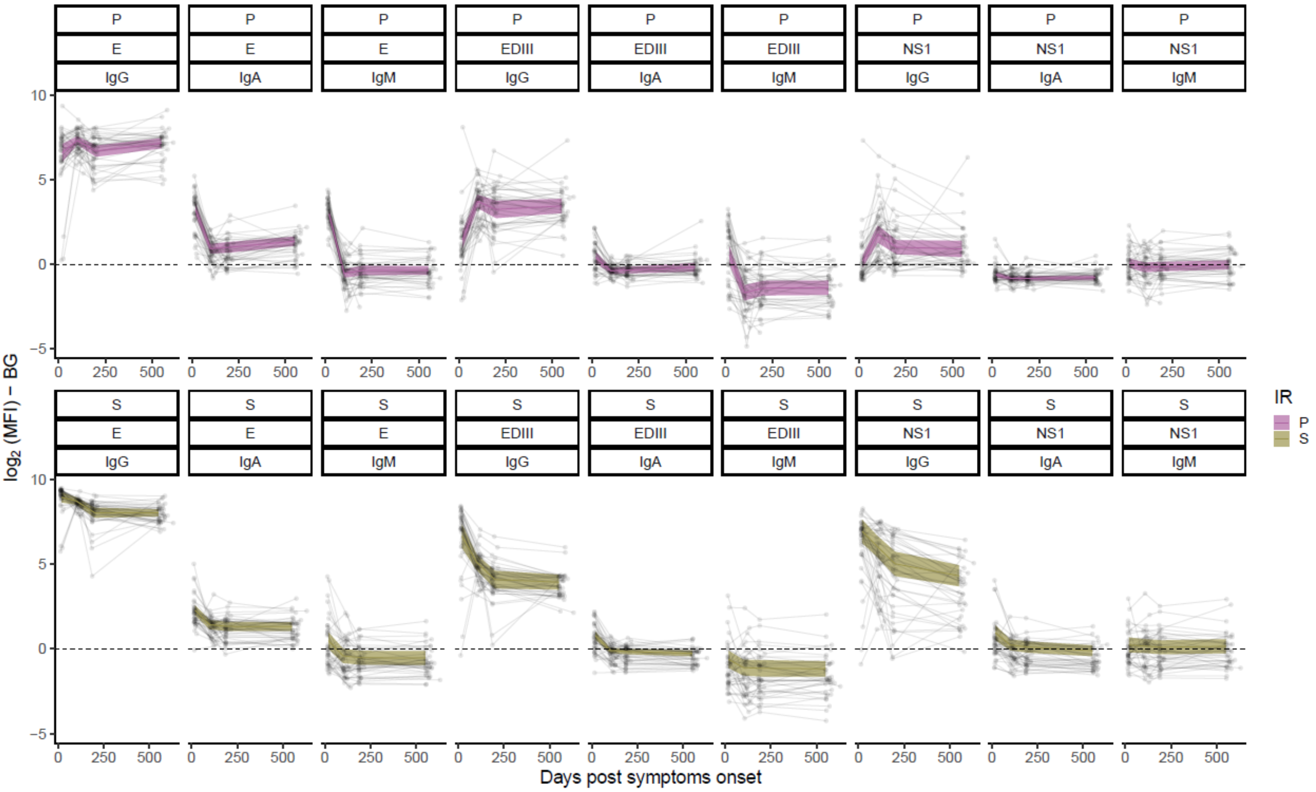


**Supplementary Figure 4. Model fit atop individual antibody trajectories for antibody responses against the cross-reactive DENV4 serotype.** The magnitude of binding antibodies was measured as $Log_2$ of mean fluorescence intensity (MFI) after background subtraction (BG; shaded area). A model was applied to estimate a linear rate of decay for three time-period segments: 30-90, >90-180, and >180-540 days post-symptom onset. Raw data on IgG, IgA, and IgM antibody levels against E, EDIII and NS1 over time for each individual with respect to antibody responses against the DENV4 serotype are shown. Model fit of the primary group is shown in pink (P) and the secondary group in beige (S), with 95% confidence intervals. The dashed line represents the background derived from DENV-naïve plasma samples.

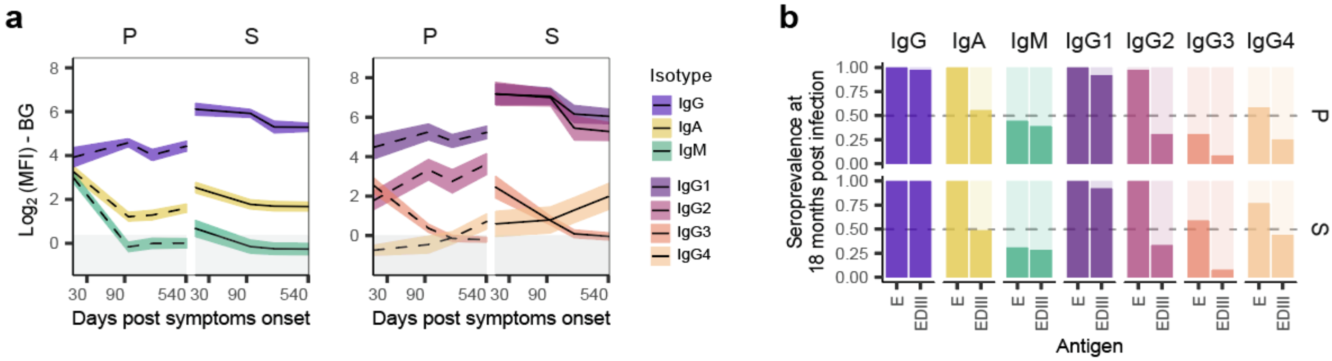


**Supplementary Figure 5. Cross-reactive antibody responses against DENV4 E protein.** Plasma antibodies against the cross-reactive DENV4 E protein were measured as $Log_2$ of mean fluorescence intensity (MFI) after background subtraction (BG; shaded area). A model was applied to estimate a linear rate of decay for three time-period segments: 30-90, >90-180, and >180-540 days post-symptom onset. **a.** Comparison of the levels DENV4 E-binding antibodies by isotype and IgG subclass after primary and secondary dengue. **b.** Seroprevalence of DENV4 E-binding antibodies in the primary versus secondary dengue group at 18 months post-symptom onset, with dashed line at 50%.

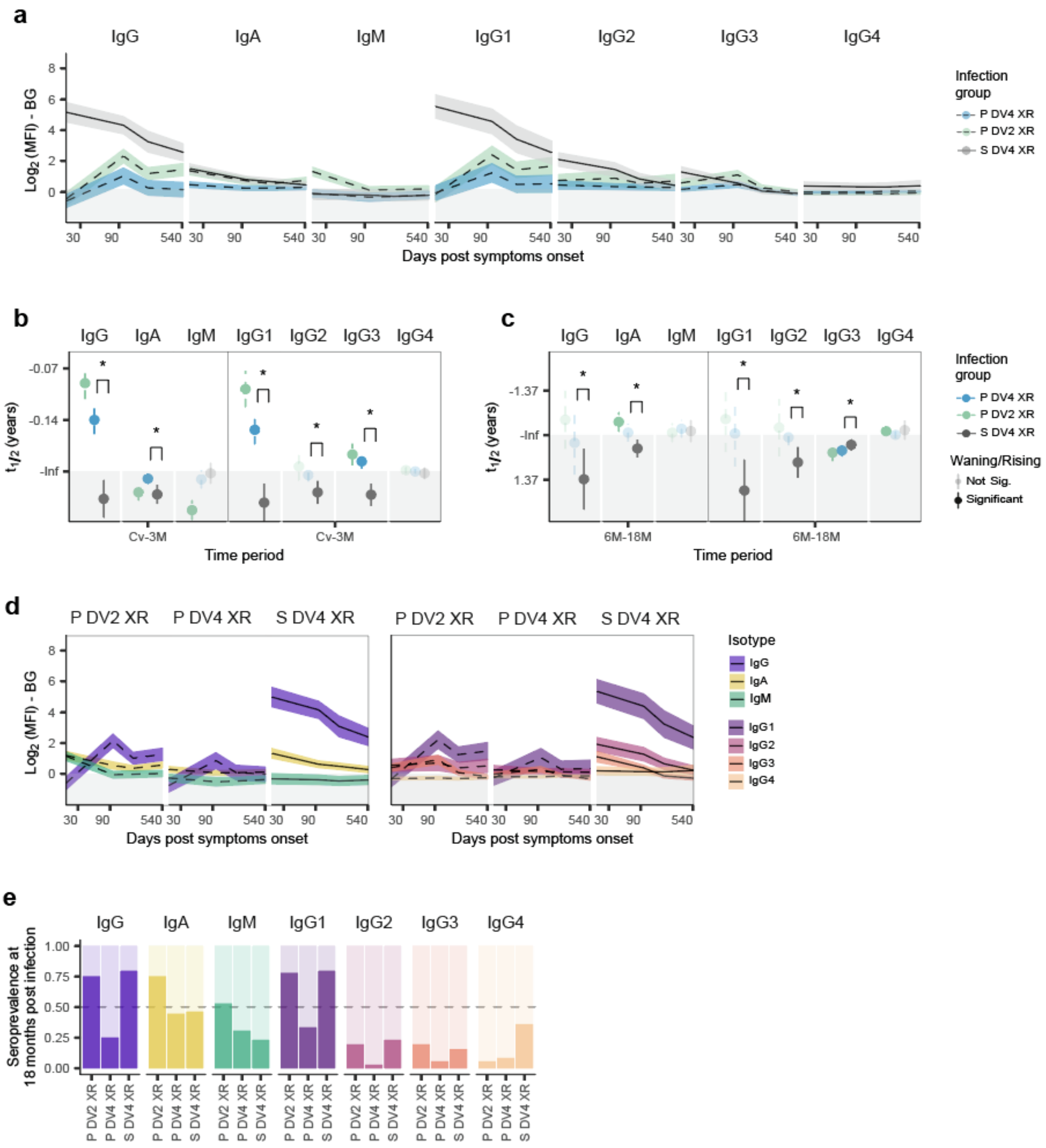


**Supplementary Figure 6. DENV serocomplex cross-reactive anti-NS1 antibodies are stable after primary dengue but wane post-secondary dengue**. Cross-reactive (XR) NS1-binding plasma antibodies from DENV1 and DENV3 infections were measured against the DENV2 and DENV4 serotypes. Binding antibodies were measured as $Log_2$ of mean fluorescence intensity (MFI) after background subtraction (BG; shaded area). A model was applied to estimate a linear rate of decay for three time-period segments: 30-90, >90-180, and >180-540 days post-symptom onset, corresponding to convalescent to 3-month (Cv-3M), 3-6 month (3-6M), and 6-18 months (6-18M) post-symptom onset. **a.** DENV2 and DENV4 XR NS1-binding antibody levels over time of primary (P; dotted line, blue and green) versus secondary (S; solid line, grey) dengue cases with 95% confidence interval for each isotype and IgG subclass. **b.** Short-term antibody half-lives ($t_{1/2}$) estimated from Cv-3M antibody decay rates, and **(c)** long-term $t_{1/2}$ estimated from 6-18M antibody decay rates. Transparent colors indicate stable decay rate with non-estimable infinite (-Inf) $t_{1/2}$ (i.e., Not significant). Solid colors are significantly rising (-) or waning (+; shaded area) antibody kinetics. Significant differences in primary versus secondary decay rates are shown with * for p<0.05. **d.** Comparison of the levels of DENV2 and DENV4 XR NS1-binding antibodies by isotype and IgG subclass after primary and secondary dengue. **e.** Seroprevalence of XR E-binding antibodies in the primary versus secondary dengue group at 18 months post-symptom onset, with dashed line at 50%.

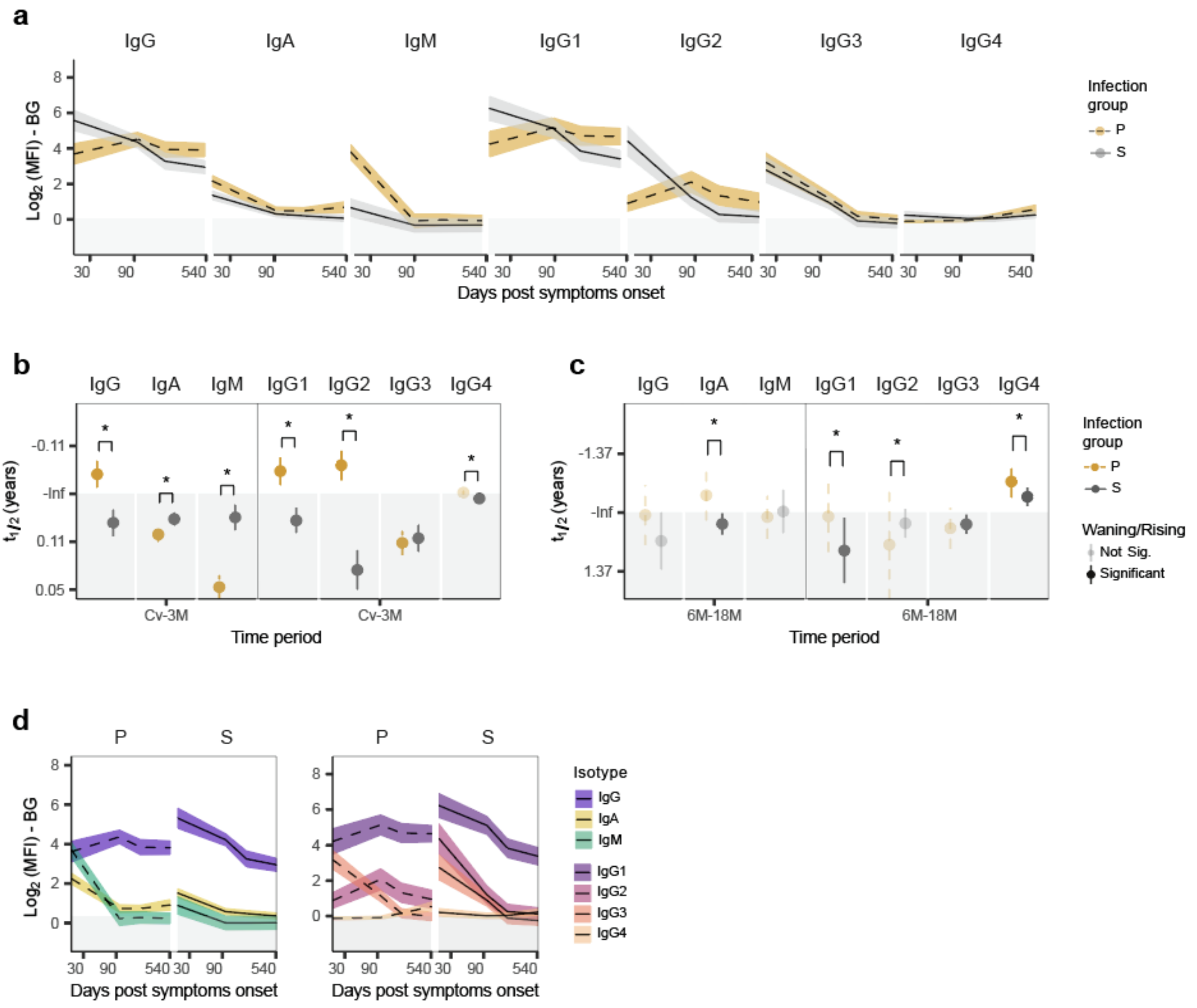


**Supplementary Figure 7. Antibody responses against homologous EDIII protein after primary and secondary dengue.** EDIII-binding plasma antibodies against the homologous infecting serotype of either DENV1 or DENV3 were measured as $Log_2$ of mean fluorescence intensity (MFI) after background subtraction (BG; shaded area). A model was applied to estimate a linear rate of decay for three time-period segments: 30-90, >90-180, and >180-540 days post-symptom onset, corresponding to convalescent to 3-month (Cv-3M), 3-6 month (3-6M), and 6-18 months (6-18M) post-symptom onset. **a.** Homologous EDIII-binding antibody levels over time of primary (P; dotted line, yellow) versus secondary (S; solid line, grey) dengue cases with 95% confidence interval for each isotype and IgG subclass. **b.** Short-term antibody half-lives ($t_{1/2}$) estimated from Cv-3M antibody decay rates, and **(c)** long-term $t_{1/2}$ estimated from 6-18M antibody decay rates. Transparent colors indicate stable decay rate with non-estimable infinite (-Inf) $t_{1/2}$ (i.e., Not significant). Solid colors are significantly rising (-) or waning (+; shaded area) antibody kinetics. Significant differences in primary versus secondary decay rates are shown with * for $p<0.05$. **d.** Comparison of the levels of homologous EDIII-binding antibodies by isotype and IgG subclass after primary and secondary dengue.

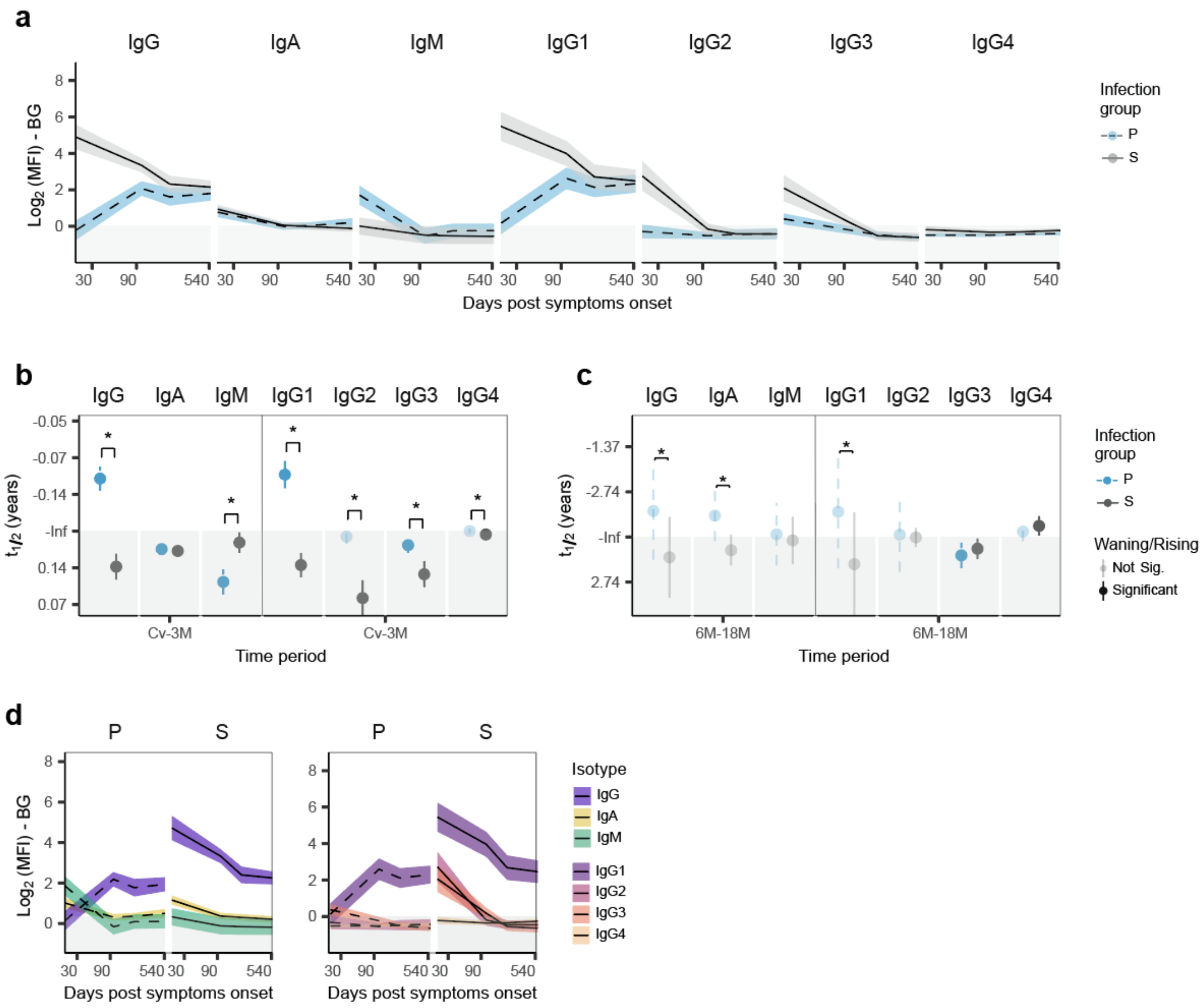


**Supplementary Figure 8. Antibody responses against cross-reactive EDIII protein after primary and secondary dengue.** Cross-reactive EDIII-binding plasma antibodies against the DENV4 serotype were measured as $Log_2$ of mean fluorescence intensity (MFI) after background subtraction (BG; shaded area). A model was applied to estimate a linear rate of decay for three time -period segments: 30-90, >90-180, and >180-540 days post-symptom onset, corresponding to convalescent to 3-month (Cv-3M), 3-6 month (3-6M), and 6-18 months (6-18M) post-symptom onset. **a.** Cross-reactive EDIII-binding antibody levels over time of primary (P; dotted line, yellow) versus secondary (S; solid line, grey) dengue cases with 95% confidence interval for each isotype and IgG subclass. **b.** Short-term antibody half-lives ($t_{1/2}$) estimated from Cv-3M antibody decay rates, and **(c)** long-term $t_{1/2}$ estimated from 6-18M antibody decay rates. Transparent colors indicate stable decay rate with non-estimable infinite (-Inf) $t_{1/2}$ (i.e., Not significant). Solid colors are significantly rising (-) or waning (+; shaded area) antibody kinetics. Significant differences in primary versus secondary decay rates are shown with * for p<0.05. **d.** Comparison of the levels of cross-reactive EDIII-binding antibodies by isotype and IgG subclass after primary and secondary dengue.

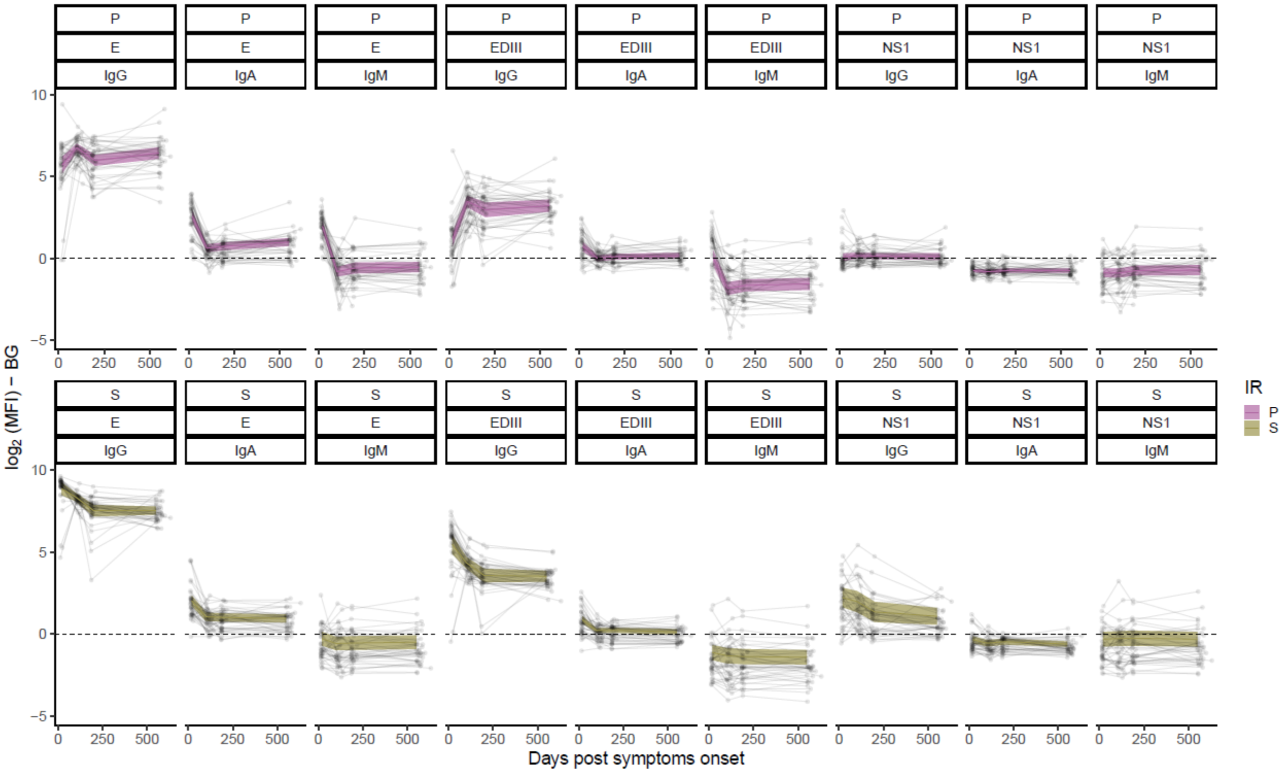


**Supplementary Figure 9. Model fit atop individual antibody trajectories for cross-reactive antibody responses against ZIKV.** The magnitude of binding antibodies was measured as $Log_2$ of mean fluorescence intensity (MFI) after background subtraction (BG; shaded area). A model was applied to estimate a linear rate of decay for three time-period segments: 30-90, >90-180, and >180-540 days post-symptom onset. Raw data on IgG, IgA, and IgM antibody levels against E, EDIII and NS1 over time for each individual with respect to antibody responses against ZIKV are shown. Model fit of the primary group is shown in pink (P) and the secondary group in beige (S), with 95% confidence intervals. Dashed line represents the background derived from DENV-naïve plasma samples.

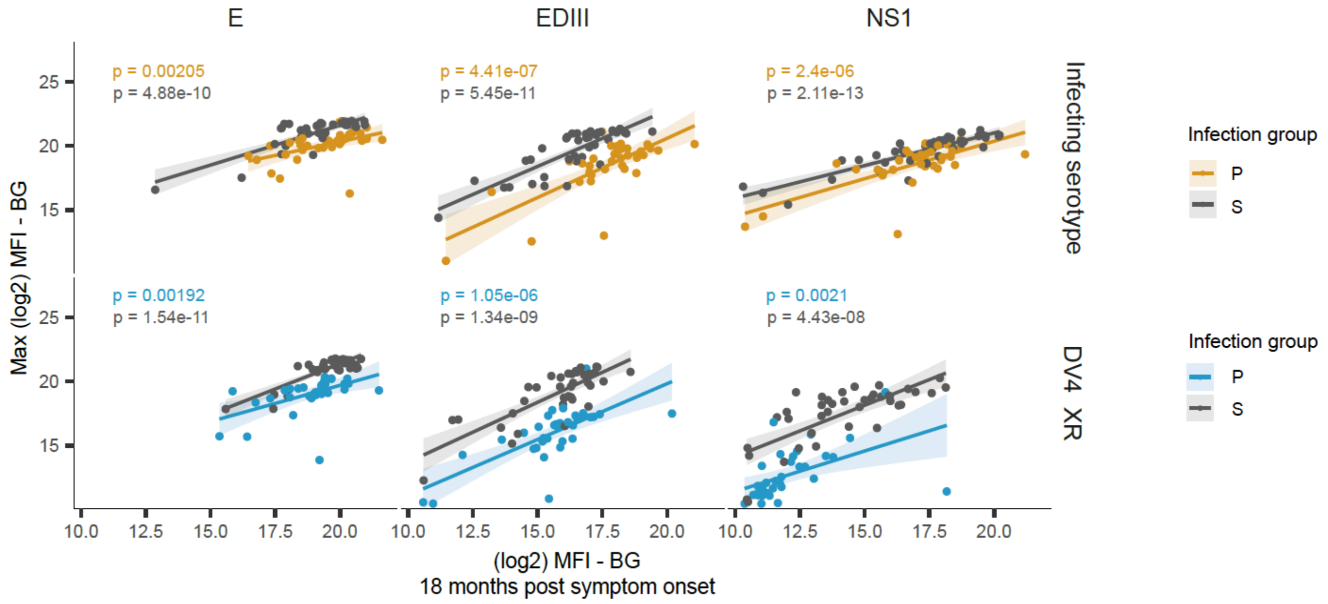


**Supplementary Figure 10. Relationship of peak early IgG1 antibody response with the level remaining at 18 months post-infection by antigen.** E-binding plasma IgG1 antibodies were measured as $Log_2$ of mean fluorescence intensity (MFI) after background subtraction (BG; shaded area). Post-primary responses against the infecting serotype are in yellow, and post-primary responses against cross-reactive DENV4 serotype are in blue (DV4_XR). All post-secondary antibody responses are in grey. The peak level of DENV-binding IgG1 (i.e., Max) was defined for each individual within 3 months of infection. The magnitude of the peak early DENV-binding IgG1 was correlated with the magnitude of the same antibody subset at 18 months post-symptom onset. The p-value of each correlation is shown in the plot. Each dot represents an individual. Axes were scaled to data for each antibody subset.

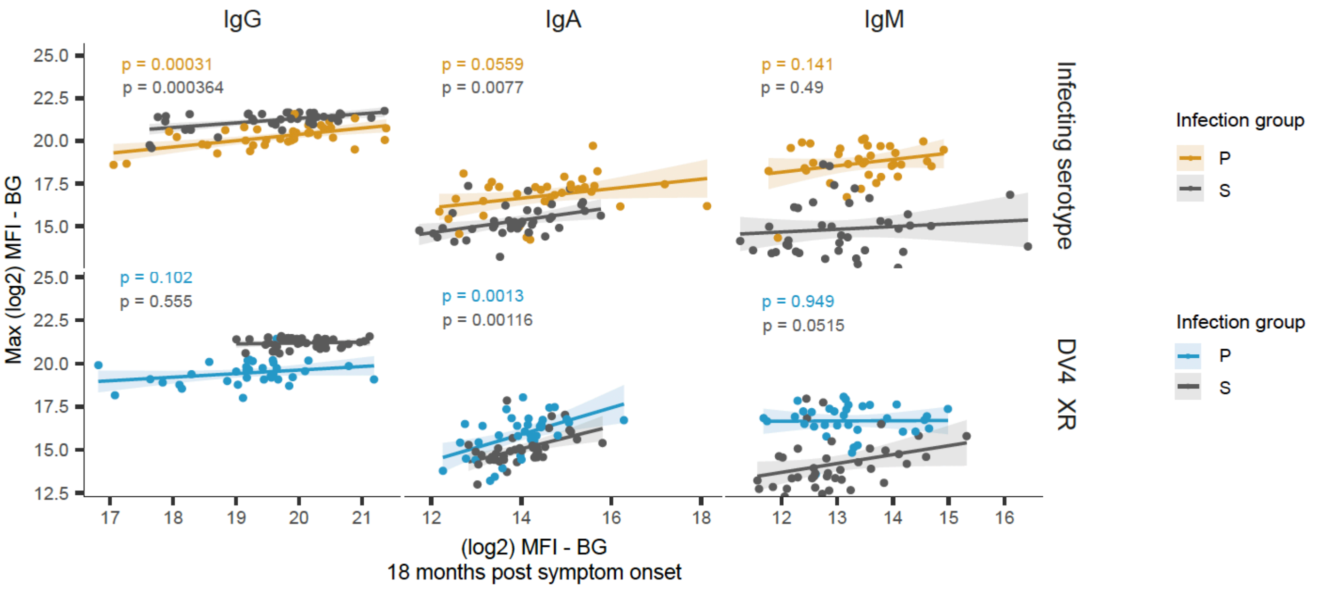


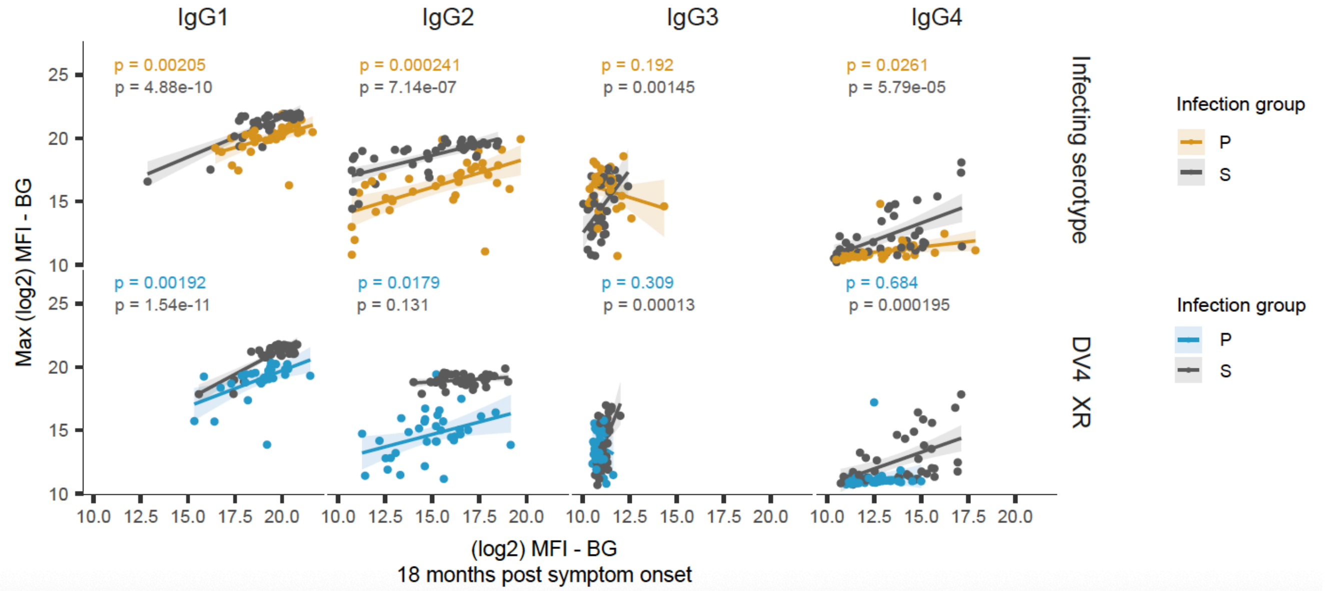


**Supplementary Figure 11. High early antibody response against E protein correlates with high antibody levels at 18 months post-infection for many antibody subsets.** E-binding plasma antibodies were measured as $Log_2$ of mean fluorescence intensity (MFI) after background subtraction (BG; shaded area). Post-primary responses against the infecting serotype are in yellow, and post-primary responses against cross-reactive DENV4 serotype are in blue (DV4_XR). All post-secondary antibody responses are in grey. The peak level of DENV-binding antibody (i.e., Max) was defined for each individual within 3 months of infection. The magnitude of the peak early DENV-binding IgG1 was correlated with the magnitude at 18 months post-symptom onset. The p-value of each correlation is shown on the plot. Each dot represents an individual. Axes were scaled to data for each antibody subset.
